# Evolution of COVID-19 patients treated with ImmunoFormulation, a combination of nutraceuticals to reduce symptomatology and improve prognosis: a multi-centred, retrospective cohort study

**DOI:** 10.1101/2020.12.11.20246561

**Authors:** Mariana Diaz Hernández, Jully Urrea, Luciano Bascoy

## Abstract

Although a vast knowledge has already been gathered on the pathophysiology of COVID-19, there are still limited, non-optimal treatment options. In this paper, we describe a multicentre, retrospective, observational study to describe the course of SARS-CoV-2 disease in patients treated with ImmunoFormulation (IF), an add-on therapy developed to decrease duration of clinical symptoms. In parallel, a group of patients that did not receive IF was used for comparison (using standard of care treatment). A total of 39 patients were evaluated. Throughout the observational period, 90% of patients recovered in the IF cohort and 47.4% in the Control cohort (p=0.0057). From the symptoms with statistically significant differences, the duration of symptoms (i.e., the time to recover from it) was shorter in the IF cohort than in control cohort (in days, average), especially for fever (2.25 x 21.78), dry cough (4.38 x 24.00), dyspnoea (3.67 x 20.00), headache (2.00 x 26.50), diarrhoea (5.25 x 25.25), and weakness (1.92 x 23.30). This demonstrates a potential promising role of IF as adjuvant therapy on the evolution of symptomatology to COVID-19 patients.

## Introduction

The coronavirus disease (COVID-19), caused by the severe acute respiratory syndrome coronavirus 2 (SARS-CoV-2), has being spread worldwide for more than 1 year. Although a vast knowledge has been gathered throughout this period, there are still limited, non-optimal treatment options. In this context, agents that can act on prophylaxis or as adjuvants to the therapies are of high value. When it comes to the pathological mechanisms of the SARS-CoV-2, it is now clear the major involvement of the immune system with consequent (hyper)inflammatory effects. In fact, some authors consider that the disease presents itself in three stages: (I) Mild (early infection, viremia phase), (II) Moderate (Pulmonary Involvement with and without Hypoxia; pneumonia phase, inflammation in the lung), and (III) Severe (Systemic Hyperinflammation) or Recovery phase.^1^ In general, the three main findings common to all phases are lymphopenia (T-cell and, more specifically, CD8^+^ T cells), imbalance between Th1 and Th2 responses (leading to cytokine storm and inflammasome activation), and decreased circulating eosinophil numbers.^2^

CD8^+^ lymphopenia (with raised C-reactive protein, D-dimer and ferritin) has been linked to the severe progression of the disease,^3,4^ and it is shown to be reversible after patient recovery, notably for mild cases^5^. When we consider previously known coronaviruses such as SARS-CoV-1 and MERS-CoV, it is also understood that T cell immunity can play a decisive role in recovery and long-term protection of patients.^6^ In addition, it seems that T cell-mediated immune response is paramount for a good prognosis, as antibody responses in coronaviruses (SARS-CoV-1) are short-lived and can even aggravate lung pathology.^6–8^ This reduction in T cells subsets are also reported to be followed by an exhaustion of effector T cells, which contributes to the defective immune response against the virus.^9,10^

The dysfunctional immune response related to the reduced functional diversity of T cells in peripheral blood is also a key parameter to predict severity, as ICU/Stage III patients tend to show a more marked Th2 profile.^3,11^ This triggers a cytokine storm which, in turn, leads to inflammatory cell infiltration and consequent secretion of proteases and reactive oxygen species (oxidative stress), which altogether contribute to the lung damage and COVID-19 severity.^12–14^ This raise in inflammatory cytokines can be observed in peripheral blood^15^, as well as a reduced level of IFN-γ,^3^ which is currently linked to a faster resolution of the infection.^16^

This knowledge brings up the concept of three points of action for improvement of symptomatology and faster recovery: regulation of the immune system, decrease of hyperinflammation and decrease of oxidative stress. Some treatments target on those have been already described and tested, but we focus here on a blend of ingredients that were first described by Ferreira *et al*.^17^ and with positive responses in isolated patients.^18^ This blend (further referred to as ImunoFormulation, IF) can potentially play a role in the prevention and/or support treatment of the symptomatology associated with COVID-19. The IF consist of: transfer factors (oligo- and polypeptides from porcine spleen, ultrafiltered at <10 kDa – Imuno TF^®^) 100 mg, 800 mg anti-inflammatory natural blend (*Uncaria tomentosa, Endopleura uchi* and *Haematoccocus pluvialis* - Miodesin™), 60 mg zinc orotate, 48 mg selenium yeast (equivalent to 96 μg of Se), 20,000 IU cholecalciferol, 300 mg ascorbic acid, 480 mg ferulic acid, 90 mg resveratrol, 800 mg spirulina, 560 mg N-acetylcysteine, 610 mg glucosamine sulphate potassium chloride, and 400 mg maltodextrin-stabilized orthosilicic acid (equivalent to 6 mg of Si – SiliciuMax^®^). The quantities correspond to the daily intake of the IF, which can be split into 3 doses, taken every 8 hours.

Thus, given the lack of gold-standard treatments, the knowledge on the virus mechanisms, and the theoretical potential benefit of the above referred adjuvant therapy, we have clinically evaluated the added value of IF for mild cases of COVID-19. In this preliminary report, we describe the course of SARS-CoV-2 disease in the patients that did or did not receive IF, based on the duration of clinical symptoms, as the basis for future clinical trials.

## METHODS

### Study design

This is a multicentre, retrospective, observational study to describe the course of SARS-CoV-2 disease in patients treated with IF. In parallel, a group of patients that did not receive IF during the course of the SARS-CoV-2 disease was used for comparison. All patients attended either one of two private clinics (Clinic Bascoy and Clínica Arvila Magna, Barcelona, Spain) from March to May 2020. Data were collected from medical registers from 02 July 2020 to 29 September 2020. Ethical approval for was granted by the Medicinal Product Research Ethics Committee of Hospital de Mar. All necessary patient/participant consent has been obtained and the appropriate institutional forms have been archived.

Secondary objectives were: (i) to describe the profile of patients (age, sex, comorbidities, concomitant medications and potential risk factors for contagion); (ii) to describe the course of SARS-CoV-2 disease in patients treated or not with IF based on the presence of symptoms at the time of the visit, two weeks and one month after the first visit for symptoms of the disease; (iii) to describe the course of SARS-CoV-2 disease in patients treated or not with IF based on the severity of the symptoms at the time of the visit, two weeks and one month after the first visit for symptoms of the disease; and (iv) to describe the adverse reactions (serious and non-serious) recorded in the patients’ medical records during treatment with IF.

### Study population

It was planned to collect data from approximately 40 patients who had tested positive in a diagnostic test for SARS-CoV-2: 20 patients who have had treated with IF and these results were compared with 20 patients who had received standard care only. Both cohorts were included without restrictions on the adjuvant treatment received.

All patients who met the screening criteria and gave their informed consent to participate were included consecutively. Inclusion criteria: patients aged 18 years or older; patients who give written informed consent to participate in the study; patients who have consulted their physician for symptoms associated with SARS-CoV-2 infection between March 2020 and May 2020; patients who have tested positive in a diagnostic test for SARS-CoV-2; patients with onset of COVID-19 symptoms ≥ 5 days prior to diagnosis of SARS-CoV-2; patients with data in the medical record from the first visit due to disease symptoms until recovery, or at least 1 month of follow-up of symptoms, whichever occurs first. Exclusion criteria included: any medical or psychological condition that, in the physician’s opinion, could compromise the patient’s ability to give informed consent; patients requiring hospital admission due to the disease.

Sample size calculation was established according to the ICH guidelines, where it was specified that the number of patients should be sufficient to provide a safe response about the issues raised. According to Lechien et al. (2020)^19^, the mean duration of mild/moderate symptoms of COVID-19 was 11.5 ± 5.7 days. A sample of 18 patients would be sufficient to estimate, with a 95% confidence and a precision of ± 2.8 days, a mean duration of symptoms with a standard deviation of 5.7 days. Assuming a loss of 10% of patients, the sample size was 20 patients. The calculations were performed with the help of the PASS package, version 2011.

### Data processing and debugging

Study data were collected in a CRD and inserted the data in a database specifically designed for the study. The database included internal consistency ranges and rules to ensure data quality control. Data recorded during the study were checked. If incomplete responses or abnormal values were seen, a query was issued to the investigator to resolve the discrepancy. When all data have been recorded and all discrepancies resolved, the database was locked the analysis was performed by the statistics department.

### Data analysis and statistical tests

The analyses of the primary and secondary objectives were performed from a single evaluable patient sample, including all patients meeting the inclusion criteria and none of the exclusion criteria. This sample of evaluable patients (EVAL set) for the description of the course of the disease was also be used for the description of the sample and the variables. Safety analyses of the secondary objective were performed on patients who have signed the informed consent (SAF set).

For comparisons between periods of continuous variables, parametric (Student’s t test for paired data) and non-parametric (Wilcoxon) tests were used, as appropriate, according to the characteristics of the study variables (assumption of normality), while categorical variables were compared using the McNemar test. The statistical tests used for comparison of the variables depended on the nature of the latter and based on the characteristics of the study variables and the number of groups to compare. The comparison between groups of quantitative variables were made using parametric (Student’s t or ANOVA) or non-parametric tests (Mann-Whitney or Kruskal-Wallis); comparison between groups of qualitative variables were made using chi-square test or Fisher test. Statistical significance level of 0.05 was used for all statistical tests. All calculations were performed using the SAS statistical software package, version 9.4.

## RESULTS

A total of 40 patients were recruited (20 in the ImmunoFormulation cohort / 20 in the Control cohort). Finally, 39 patients were Evaluable (EVAL set) (20 ImmunoFormulation cohort / 19 Control cohort) for efficacy variables (Figure 1). The control cohort received standard care only, while the ImmunoFormulation cohort received standard care and the IF, prepared by a local compounding pharmacy (concomitant medication during the observational study is summarized in Table S1). The median time between the first consultation for symptomatology and positive diagnosis test for SARS-CoV-2 was 6.00 days in the IF and 15.00 days in the Control cohort, observing statistically significant differences (p=0.0004) (Table 1, which also describes the population’s sociodemographic data and the profile of comorbidities).

**Table 1.** Patient’s characteristics upon the first consultation for symptomatology associated with SARS-CoV-2 infection.

| **Characteristics ImmunoFormulation** | **ImmunoFormulation cohort (n=20)** |
| --- | --- |
| Age, year [mean (SD)] * | 54.25 (14.24) |
| Male gender, n (%) | 9 (45.0) |
| Time between first consultation for symptomatology and positive diagnosis for SARS-CoV-2, days [median (25; P75)] * | 6.00 (5.00; 6.00) |
| Time between the start date of the first symptom and start date of the ImmunoFormulation treatment, days [median (25; P75)] | 6.00 (5.00; 6.00) |
| **Type of patient according to the most common severity first symptom<sup>a</sup>** |  |
| Predominance of mild severity, n (%) | 10 (50.0) |
| Predominance of moderate severity, n (%) | 6 (30.0) |
| Predominance of severe severity, n (%) | 4 (20.0) |
| **Comorbidities\* (n= 8 patients with 14 relevant comorbidities)** |  |
| Cardiac disorders, n (%) | 0 (0.0) |
| Vascular disorders, n (%) | 4 (28.6) |
| Endocrine disorders, n (%) | 1 (7.1) |
| Musculoskeletal disorders, n (%) | 1 (7.1) |
| Neoplasms, n (%) | 3 (21.4) |
| Nervous system disorders, n (%) | 0 (0.0) |
| Psychiatric disorders, n (%) | 0 (0.0) |
| Renal disorders, n (%) | 0 (0.0) |
| Respiratory disorders, n (%) | 2 (14.3) |
| **Characteristics Control** | **Control cohort (n=19)** |
| Age, year [mean (SD)] * | 81.16 (11.30) |
| Male gender, n (%) | 3 (15.8) |
| Time between first consultation for symptomatology and positive diagnosis for SARS-CoV-2, days [median (25; P75)] * | 15.00 (6.00; 21.00) |
| Time between the start date of the first symptom and start date of the ImmunoFormulation treatment, days [median (25; P75)] | - |
| **Type of patient according to the most common severity first symptom<sup>a</sup>** |  |
| Predominance of mild severity, n (%) | 7 (36.8) |
| Predominance of moderate severity, n (%) | 5 (26.3) |
| Predominance of severe severity, n (%) | 7 (36.3) |
| **Comorbidities\*\* (n= 19 patients with 52 relevant comorbidities)** |  |
| Cardiac disorders, n (%) | 11 (21.2) |
| Vascular disorders, n (%) | 14 (26.9) |
| Endocrine disorders, n (%) | 0 (0.0) |
| Musculoskeletal disorders, n (%) | 4 (7.7) |
| Neoplasms, n (%) | 0 (0.0) |
| Nervous system disorders, n (%) | 1 (1.9) |
| Psychiatric disorders, n (%) | 5 (9.6) |
| Renal disorders, n (%) | 2 (3.8) |
| Respiratory disorders, n (%) | 4 (7.7) |

<sup>a</sup>A single patient could have more than one symptom. In this case, the most common first symptom was analysed. When groups would be directly compared, characteristics indicated by asterisk are statistically different ( $p < 0.05$ ).

**Figure 1.**
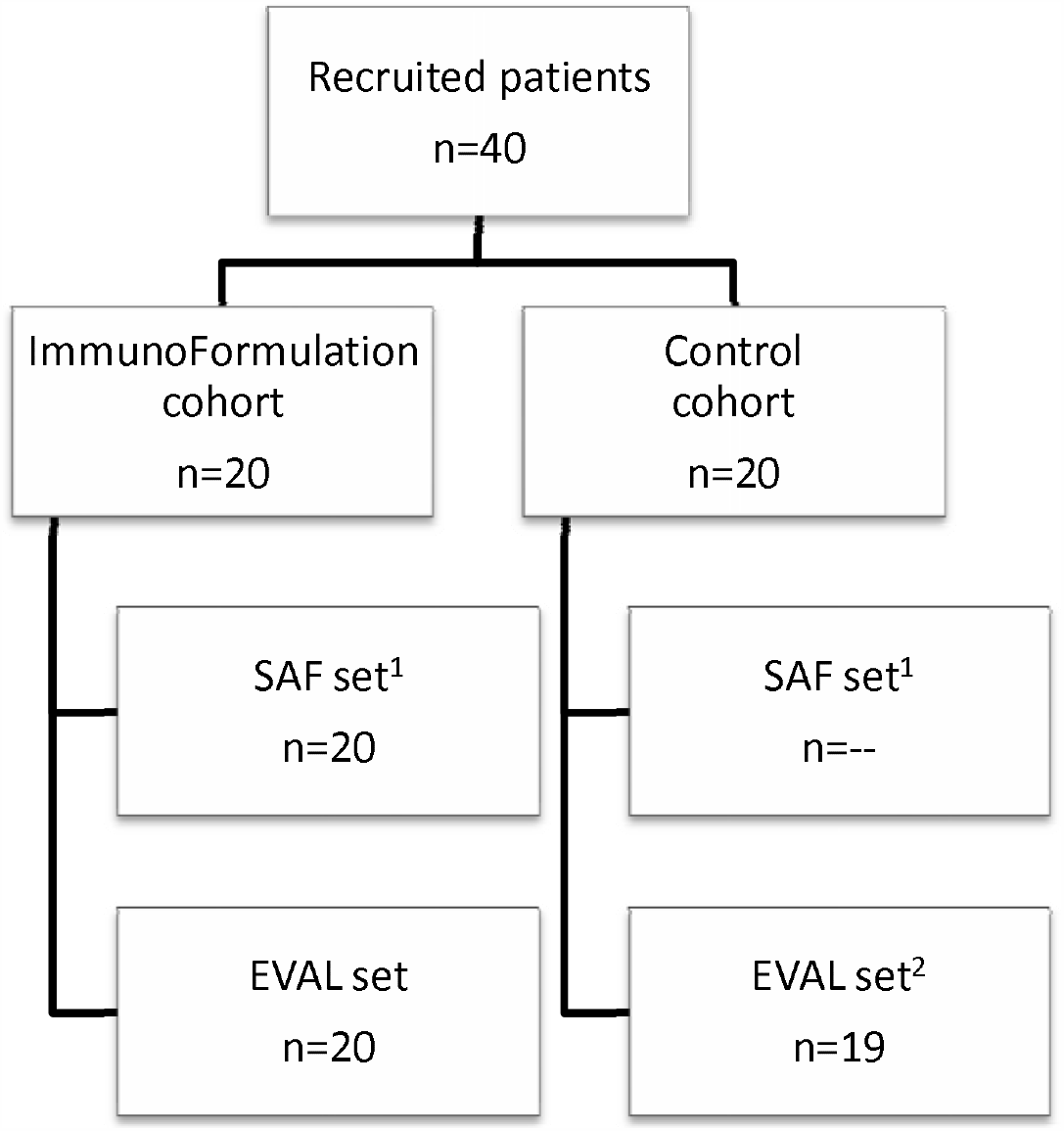
Flow chart of the study. ^1^ Description of adverse reactions only were analysed in the ImmunoFormulation cohort. ^2^ One patient did not meet Inclusion Criterion (patients with onset of COVID-19 symptoms ≥ 5 days prior to diagnosis of SARS-CoV-2). SAF = Safety. EVAL = Evaluable patients.

Overall, most common first symptoms were (Table S2): weakness (53.8%), fever (51.3%), dry cough (41.0%), dyspnoea (30.8%) and headache (17.9%). In the ImmunoFormulation cohort the most common first symptoms were: weakness (60.0%), headache (30.0%), abdominal pain (20.0%) and general discomfort (20.0%). In the Control cohort the most common first symptoms were: fever (89.5%), dry cough (68.4%), dyspnoea (57.9%), weakness (47.4%) and hypoxemia (21.1%). Statistically significant differences between cohorts were observed in fever (p<0.0001), dry cough (p=0.0011), dyspnoea (p=0.0004) and hypoxemia (p=0.0471). Patients were classified for the severity of their first severity symptoms according to the most common first symptoms: In the ImmunoFormulation cohort the 50.0% of the patients were classified as mild, 30.0% as moderate and 20.0% as severe (Table 1). In the Control cohort 36.8% of the patients were classified as mild, 26.3% as moderate and 36.8% as severe. No statistically significant differences between cohorts were observed (p=0.4927). Detailed information on each symptom can be found in Table S3.

### Primary outcomes

Throughout the observational period, 90% of patients recovered in the ImmunoFormulation cohort and 47.4% in the Control cohort (p=0.0057) (Figure 2). According to the most severe first symptoms, in the ImmunoFormulation cohort, the mean (SD) days with some symptoms from the start of IF treatment to the end of the observational period was 11.22 (10.06) days in mild symptoms, 17.57 (8.36) days in moderate symptoms and 16.00 (8.76) days in severe symptoms. In the Control cohort, the mean (SD) days with some symptoms to the end of the observational period (end observational period - start first symptom) was 28.00 (4.47) days in mild symptoms, 28.00 (4.47) days in moderate symptoms and 25.42 (5.52) days in severe symptoms (Table S4).

**Figure 2.**
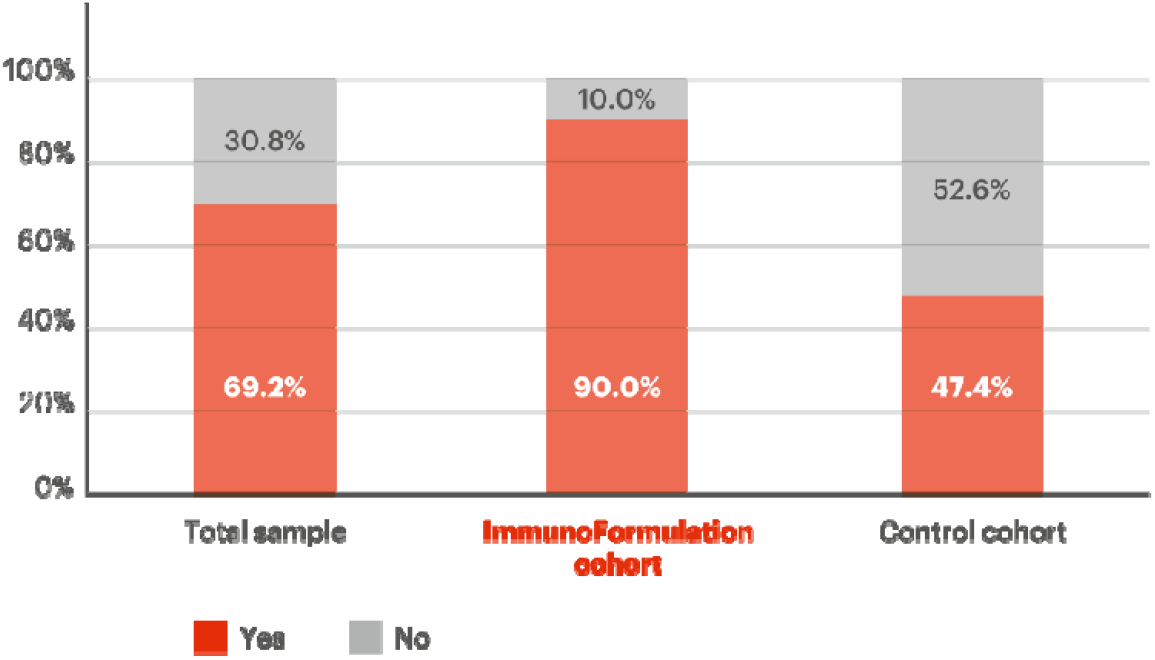
Patients recovered from start of the first symptom to the end of the observational period.

The duration of symptoms in both cohorts (time to recover from start of the first symptom), as well as the percentage of recovery of each symptom by the end of the observational period is described in Table 2. From the symptoms with statistically significant differences, the duration of symptoms (i.e., the time to recover from it) was shorter in the ImmunoFormulation cohort, especially for fever, headache, and weakness, which ended in less than 2 days.

**Table 2.** Total recovery duration of symptoms associated with COVID-19 stratified by the most common symptoms.

| Symptom | ImmunoFormulation cohort |  | Control cohort |
| --- | --- | --- | --- |
|  | Time to recover from start of the first symptom / patients recovered by the end of the observational period | Time to recover from start of treatment | Time to recover from start of the first symptom / patients recovered by the end of the observational period |
| Fever<sup>*</sup> | 3.35 (2.87) / 100.0% | 2.25 (0.91) / 100.0% | 21.78 (7.75) / 66.7% |
| Dry cough<sup>*</sup> | 6.15 (6.52) / 100.0% | 4.38 (6.31) / 100.0% | 24.00 (7.39) / 53.3% |
| Dyspnoea<sup>*</sup> | 5.67 (6.35) / 100.0% | 3.67 (2.08) / 100.0% | 20.00 (7.29) / 71.4% |
| Loss of taste and smell | 21.55 (7.27) / 90.9% | 19.73 (4.67) / 90.9% | 26.50 (4.95) / 0.0% |
| Headache<sup>*</sup> | 6.25 (1.98) / 100.0% | 2.00 (1.31) / 100.0% | 26.50 (4.95) / 0.0% |
| Diarrhoea<sup>*</sup> | 8.75 (4.35) / 100.0% | 5.25 (5.85) / 100.0% | 25.25 (3.20) / 25.0% |
| Weakness<sup>*</sup> | 7.42 (1.08) / 100.0% | 1.92 (0.67) / 100.0% | 23.30 (9.37) / 50.0% |

<sup>\*</sup> p < 0.05. Only symptoms present in more than 2 patients in each group are shown (full overview is shown in Tables S5 and S6).

As for the adverse reactions’ evaluation, no patient presented adverse drug reactions (Table 3).

**Table 3.** Adverse drug reactions in the ImmunoFormulation cohort.

| **Patients with adverse drug reactions** | **ImmunoFormulation cohort, n (%)** |
| --- | --- |
| *Adverse reactions patients (ADR)* | 20 (100.0%) |
| Patients with adverse drug reactions | 0 (0.0%) |
| Patients without adverse drug reactions | 20 (100.0%) |

## DISCUSSION

The lack of standard treatment for COVID-19 creates the need for investigation of strategies that can either target SARS-CoV-2 to eliminate it or to improve the symptomatology and strengthen the natural defences. We aimed on this second option and evaluated the use of an add-on therapy described previously on literature.^17,18^ Comparing the two cohorts, a clear difference was seen in the resolution of most symptoms, including fever, dry cough, dyspnoea, headache, diarrhoea, and weakness. Overall, the reduction in time for the resolution of the symptoms indicate a possible positive effect for IF as an add-on therapy for COVID-19.

Robust studies showing the time for recovery of symptoms are still lacking, as most of them focus on the time for symptom onset and in the rate of recovery/complications. The time from exposure to symptom onset is usually reported as is in average 11.5 days, and the time between symptom onset and hospital admission about 7 days.^20,21^ Usually the first symptoms (Stage I: fever, dry cough, headache, diarrhoea) appear between 0 to 4 days; the Stage II symptoms (hypoxia) in 5-13 days; and Stage III symptoms (ARDS, cardiac failure, shock) after 14 days of infection.^1^ This is in concordance with what was found by Wang *et al*, a median 5 days (range 2-8 days) for the progression from mild-moderate cases to severe condition, and a hospital stay range from 14 to 22 days.^22^

As an attempt for comparison, Carfi *et al*.^23^ evaluated a population similar to our IF cohort in sociodemographic terms: patients with mean age of 56.5 (± 14.6) years, and 63% were men; the difference is that they evaluated hospitalized patients. They assessed the patients for a mean of 60.3 days after onset of the first COVID-19 symptom and observed that only 12.6% were completely free of any COVID-19–related symptom, while 32% had 1 or 2 symptoms and 55% had 3 or more. A report from Imperial College of London^24^ showed that the mean time for recovery after symptom onset is 20.51 (± 6.69) days. In contrast, 90.0% of the IF cohort of the present study recovered during the observational period (30 days), and the most common symptoms were resolved within around 2 to five days (except for loss of taste and smell, which is known to be a long-lasting or irreversible complication of COVID-19^25^).

A similar population studied was also reported by Chen *et al*.^26^: patients with mild cases, a median of 51 years, and a percentage of 50.6% men. In this study, the estimated median duration of fever was 10 days (CI: 8-11 days), after onset of symptoms – in our findings, the duration of fever was 3.35 days after the onset of symptoms and 2.25 days after the start of treatment.

Obtaining fast patient recovery is important, as the persistence of symptoms can reflect the worsening in his prognosis. For example, for severe cases, the symptoms can last for more than 28 days, leading to hyperinflammation/hypercoagulation responses and pulmonary fibrosis formation.^27^ The improvement in the time needed for recovery of the symptoms in the IF cohort can be related to the multiple mechanisms that the components of the IF theoretically acts on as described earlier by Ferreira et al.^17^ We will highlight four. First, immune system regulation. This can be related to macrophage activation by Imuno TF^®28,29^ and spirulina,^30,31^ to development of neutrophils by Spirulina and Zinc,^32^ to activation of NK-cells by Imuno TF^®^, Spirulina, Zinc, Vitamin C, and Resveratrol,^31,33–38^ to the increase in T-cells functions by Spirulina, Vitamin C and Vitamin D_3_,^32,39–41^ and to CD4^+^ cells activation by Imuno TF^®^ and Selenium, which can regulate the antigenic stimulus triggering CD4^+^ Th1 cells to produce IFN-γ, IL-1 and TNF-α.^28,29,32,42–44^ In addition, Imuno TF^®^ positively regulates Th1 cytokines, while decreases the release of Th2 cytokines (IL-4, IL-5, IL-6, IL-13).^45^ This is relevant once there is evidence that the Th2 overresponse are linked to bronchoconstriction, dyspnea and exacerbations of allergic airways diseases.^46^

Secondly, targeting the virus itself by: (i) avoiding the virus to enter the cell: N-acetylcysteine and Resveratrol have shown DPP4R inhibitory effect, and Resveratrol also showed potential to block the binding of ACE2 at the molecular level;^47,48^ and (ii) decreasing virus replication: studies have shown that high concentration of intracellular Zinc inhibited the replication of SARS coronavirus (SARS-CoV) and other RNA viruses, through inhibition of RNA polymerase.^49^ Resveratrol can act synergistically with Zinc, as it has been shown to increase the intracellular entrance of Zinc.^50^ In addition, N-acetylcysteine, Selenium, and Glucosamine might be expected to help to prevent and control RNA virus infections because they amplify the signalling functions of TLR7 and mitochondrial antiviral-signalling protein (MAVS) in type 1 IFN production.^51^ Recently, *U. tomentosa* bark extract (one of the components of Miodesin™) has shown antiviral effect against SARS-CoV-2 on Vero E6 cells.^52^

Third, the IF effects the inflammatory process generated by the infection. As Vitamin D_3_ exhibits anti-inflammatory properties, it could potentiate innate immunity while controlling the potentially harmful inflammatory response. This immunoregulatory effect could in turn prevent hyperinflammatory response caused by respiratory tract infections.^32,53–55^ In fact, vitamin D3 and Imuno TF^®^ can decrease the IL-6 effect, which is a known marker of poor outcome in critically ill patients.^45,56^ For example, the evaluation of a large number of patients from several countries has demonstrated that Vitamin D_3_ may reduce COVID-19 severity by a suppressive effect on the cytokines storm, and therefore improve clinical outcomes of patients.^57–62^ Indeed, a negative correlation between vitamin D_3_ serum levels and the number of cases of COVID-19, and also the number of deaths, has been observed.^63–65^ Vitamin C, can increase lymphocyte B and T proliferation and differentiation at a controlled rate.^40,66,67^ Resveratrol and Ferulic acid can inhibit the TLR4 signalling pathway, which provides potential protection against tissue damage (including lung) coming from excessive inflammatory response.^51^ Among the potential pharmacological effects of Ferulic acid figures the decrease of the serological concentration of TNF-α and IL-1β, the suppression in TLR4 expression and the reduced activation of MAPK and NF-κB.^68,69^ Another ingredient, Miodesin™, was shown recently to decrease inflammation through inhibition of the release of cytokines (IL-1β, IL-6, IL-8, and TNF-α) and chemokines (CCL2, CCL3, and CCL5) and the expression of NF-κB, inflammatory enzymes (COX-1, COX-2, PLA2, iNOS), and chemokines (CCL2, CCL3, and CCL5).^70^

Finally, the add-on treatment provided was idealized to also act on the oxidative stress. Phase 2 inductive nutraceuticals as Ferulic acid and Resveratrol induce various peroxidase enzymes (enzymes that neutralize hydrogen peroxidase, a reactive oxygen species) and promote synthesis of glutathione. Glutathione production can also be promoted by administration of N-acetylcysteine. Selenium supplementation might also be appropriate in this context.^51^ Besides, other nutraceuticals with antioxidant properties such as Vitamin C, Spirulina and Astaxanthin can also contribute to reduce the oxidative stress.^51,66,67,71–74^

As a limitation of our study, we can point out the differences in age of the cohorts. Therefore, we can understand the data as a description of the fast times needed to recover from the most common COVID-19 symptoms, rather than a direct comparison between the cohorts.

## CONCLUSION

This retrospective observational study demonstrates a potential promising role of ImmunoFormulation as adjuvant therapy on the evolution of symptomatology to COVID-19 patients. Specially for the symptoms fever, dry cough, dyspnoea, headache, diarrhoea and weakness, the recovery time for the treated cohort was significant shorter in comparison to the control cohort. A controlled, double-blind, randomized clinical trial in a larger population is therefore currently being conducted.

## Data Availability

Data will be available upon request.

