## Supplemental Tables for "Evolution of COVID-19 patients treated with ImmunoFormulation, a combination of nutraceuticals to reduce symptomatology and improve prognosis: a multi-centred, retrospective cohort study"

Mariana Diaz Hernández[1], Jully Urrea[1], Luciano Bascoy[2]
[1]*Clínica Arvila Magna. Avinguda Diagonal, 442, 08037 Barcelona, Spain.*
[2]*Clinic Bascoy. Carrer d'Horaci 9, 08022 Barcelona, Spain.*

### SUPPLEMENTARY MATERIAL

**Table S1. Concomitant medication by maximum degree of severity in the initial symptomatology**

| | **Total** n (%) | **Mild** n (%) | **Moderate** n (%) | **Severe** n (%) | **p**[1] |
|---|---|---|---|---|---|
| **Total sample** | | | | | |
| **Patients with concomitant medication** | **39 (100.0%)** | **11 (100.0%)** | **12 (100.0%)** | **16 (100.0%)** | 0.0353(f) |
| Yes | 34 (87.2%) | 10 (90.9%) | 8 (66.7%) | 16 (100.0%) | |
| No | 5 (12.8%) | 1 (9.1%) | 4 (33.3%) | 0 (0.0%) | |
| **N=34 patients with n=173 concomitant medications** | **173 (100.0%)** | **32 (100.0%)** | **41 (100.0%)** | **100 (100.0%)** | 0.7336 |
| AGENTS ACTING ON THE RENIN-ANGIOTENSIN SYSTEM | 8 (4.6%) | 2 (6.3%) | 3 (7.3%) | 3 (3.0%) | |
| ALL OTHER THERAPEUTIC PRODUCTS | 1 (0.6%) | 0 (0.0%) | 0 (0.0%) | 1 (1.0%) | |
| ANALGESICS | 27 (15.6%) | 10 (31.3%) | 5 (12.2%) | 12 (12.0%) | |
| ANTI-ACNE PREPARATIONS | 1 (0.6%) | 0 (0.0%) | 0 (0.0%) | 1 (1.0%) | |
| ANTIBACTERIALS FOR SYSTEMIC USE | 33 (19.1%) | 3 (9.4%) | 8 (19.5%) | 22 (22.0%) | |
| ANTIBIOTICS AND CHEMOTHERAPEUTICS FOR DERMATOLOGICAL USE | 1 (0.6%) | 0 (0.0%) | 0 (0.0%) | 1 (1.0%) | |
| ANTIDIARRHEALS. INTESTINAL ANTIINFLAMMATORY/ANTIINFECTIVE AGENTS | 17 (9.8%) | 2 (6.3%) | 7 (17.1%) | 8 (8.0%) | |
| ANTIEPILEPTICS | 1 (0.6%) | 0 (0.0%) | 0 (0.0%) | 1 (1.0%) | |
| ANTIINFLAMMATORY AND ANTIRHEUMATIC PRODUCTS | 5 (2.9%) | 0 (0.0%) | 1 (2.4%) | 4 (4.0%) | |
| ANTINEOPLASTIC AGENTS | 1 (0.6%) | 1 (3.1%) | 0 (0.0%) | 0 (0.0%) | |
| ANTITHROMBOTIC AGENTS | 3 (1.7%) | 0 (0.0%) | 0 (0.0%) | 3 (3.0%) | |
| BETA BLOCKING AGENTS | 8 (4.6%) | 0 (0.0%) | 2 (4.9%) | 6 (6.0%) | |
| CALCIUM CHANNEL BLOCKERS | 2 (1.2%) | 1 (3.1%) | 0 (0.0%) | 1 (1.0%) | |
| CORTICOSTEROIDS. DERMATOLOGICAL PREPARATIONS | 1 (0.6%) | 0 (0.0%) | 0 (0.0%) | 1 (1.0%) | |
| DIURETICS | 11 (6.4%) | 1 (3.1%) | 2 (4.9%) | 8 (8.0%) | |
| DRUGS FOR ACID RELATED DISORDERS | 4 (2.3%) | 0 (0.0%) | 1 (2.4%) | 3 (3.0%) | |
| DRUGS FOR CONSTIPATION | 4 (2.3%) | 1 (3.1%) | 0 (0.0%) | 3 (3.0%) | |
| DRUGS FOR OBSTRUCTIVE AIRWAY DISEASES | 17 (9.8%) | 5 (15.6%) | 4 (9.8%) | 8 (8.0%) | |
| DRUGS USED IN DIABETES | 9 (5.2%) | 2 (6.3%) | 2 (4.9%) | 5 (5.0%) | |

| Table S1. Concomitant medication by maximum degree of severity in the initial symptomatology | | | | | |
|---|---|---|---|---|---|
| | **Total** n (%) | **Mild** n (%) | **Moderate** n (%) | **Severe** n (%) | **p[1]** |
| ENDOCRINE THERAPY | 1 (0.6%) | 0 (0.0%) | 1 (2.4%) | 0 (0.0%) | |
| LIPID MODIFYING AGENTS | 3 (1.7%) | 1 (3.1%) | 1 (2.4%) | 1 (1.0%) | |
| OPHTHALMOLOGICALS | 1 (0.6%) | 0 (0.0%) | 0 (0.0%) | 1 (1.0%) | |
| PSYCHOANALEPTICS | 7 (4.0%) | 2 (6.3%) | 1 (2.4%) | 4 (4.0%) | |
| PSYCHOLEPTICS | 2 (1.2%) | 1 (3.1%) | 1 (2.4%) | 0 (0.0%) | |
| THYROID THERAPY | 1 (0.6%) | 0 (0.0%) | 0 (0.0%) | 1 (1.0%) | |
| UROLOGICALS | 1 (0.6%) | 0 (0.0%) | 1 (2.4%) | 0 (0.0%) | |
| VASOPROTECTIVES | 3 (1.7%) | 0 (0.0%) | 1 (2.4%) | 2 (2.0%) | |
| **ImmunoFormulation cohort** | | | | | |
| **Patients with concomitant medication** | **20 (100.0%)** | **9 (100.0%)** | **7 (100.0%)** | **4 (100.0%)** | 0.0844(f) |
| Yes | 15 (75.0%) | 8 (88.9%) | 3 (42.9%) | 4 (100.0%) | |
| No | 5 (25.0%) | 1 (11.1%) | 4 (57.1%) | 0 (0.0%) | |
| **N=15 patients with n=34 concomitant medications** | **34 (100.0%)** | **15 (100.0%)** | **6 (100.0%)** | **13 (100.0%)** | 0.5904 |
| AGENTS ACTING ON THE RENIN-ANGIOTENSIN SYSTEM | 2 (5.9%) | 1 (6.7%) | 1 (16.7%) | 0 (0.0%) | |
| ANALGESICS | 13 (38.2%) | 7 (46.7%) | 3 (50.0%) | 3 (23.1%) | |
| ANTI-ACNE PREPARATIONS | 1 (2.9%) | 0 (0.0%) | 0 (0.0%) | 1 (7.7%) | |
| ANTIBACTERIALS FOR SYSTEMIC USE | 1 (2.9%) | 0 (0.0%) | 0 (0.0%) | 1 (7.7%) | |
| ANTIBIOTICS AND CHEMOTHERAPEUTICS FOR DERMATOLOGICAL USE | 1 (2.9%) | 0 (0.0%) | 0 (0.0%) | 1 (7.7%) | |
| ANTINEOPLASTIC AGENTS | 1 (2.9%) | 1 (6.7%) | 0 (0.0%) | 0 (0.0%) | |
| ANTITHROMBOTIC AGENTS | 1 (2.9%) | 0 (0.0%) | 0 (0.0%) | 1 (7.7%) | |
| CALCIUM CHANNEL BLOCKERS | 2 (5.9%) | 1 (6.7%) | 0 (0.0%) | 1 (7.7%) | |
| CORTICOSTEROIDS. DERMATOLOGICAL PREPARATIONS | 1 (2.9%) | 0 (0.0%) | 0 (0.0%) | 1 (7.7%) | |
| DIURETICS | 1 (2.9%) | 0 (0.0%) | 0 (0.0%) | 1 (7.7%) | |
| DRUGS FOR CONSTIPATION | 2 (5.9%) | 1 (6.7%) | 0 (0.0%) | 1 (7.7%) | |
| DRUGS FOR OBSTRUCTIVE AIRWAY DISEASES | 3 (8.8%) | 3 (20.0%) | 0 (0.0%) | 0 (0.0%) | |
| ENDOCRINE THERAPY | 1 (2.9%) | 0 (0.0%) | 1 (16.7%) | 0 (0.0%) | |
| LIPID MODIFYING AGENTS | 3 (8.8%) | 1 (6.7%) | 1 (16.7%) | 1 (7.7%) | |
| THYROID THERAPY | 1 (2.9%) | 0 (0.0%) | 0 (0.0%) | 1 (7.7%) | |
| **Control cohort** | | | | | |
| **Patients with concomitant medication** | **19 (100.0%)** | **2 (100.0%)** | **5 (100.0%)** | **12 (100.0%)** | -- |
| Yes | 19 (100.0%) | 2 (100.0%) | 5 (100.0%) | 12 (100.0%) | |
| No | 0 (0.0%) | 0 (0.0%) | 0 (0.0%) | 0 (0.0%) | |
| **N=19 patients with n=139 concomitant medications** | **139 (100.0%)** | **17 (100.0%)** | **35 (100.0%)** | **87 (100.0%)** | 0.9656 |
| AGENTS ACTING ON THE RENIN-ANGIOTENSIN SYSTEM | 6 (4.3%) | 1 (5.9%) | 2 (5.7%) | 3 (3.4%) | |
| ALL OTHER THERAPEUTIC PRODUCTS | 1 (0.7%) | 0 (0.0%) | 0 (0.0%) | 1 (1.1%) | |

**Table S1. Concomitant medication by maximum degree of severity in the initial symptomatology**

| | Total n (%) | Mild n (%) | Moderate n (%) | Severe n (%) | p[1] |
|---|---|---|---|---|---|
| ANALGESICS | 14 (10.1%) | 3 (17.6%) | 2 (5.7%) | 9 (10.3%) | |
| ANTIBACTERIALS FOR SYSTEMIC USE | 32 (23.0%) | 3 (17.6%) | 8 (22.9%) | 21 (24.1%) | |
| ANTIDIARRHEALS. INTESTINAL ANTIINFLAMMATORY/ANTIINFECTIVE AGENTS | 17 (12.2%) | 2 (11.8%) | 7 (20.0%) | 8 (9.2%) | |
| ANTIEPILEPTICS | 1 (0.7%) | 0 (0.0%) | 0 (0.0%) | 1 (1.1%) | |
| ANTIINFLAMMATORY AND ANTIRHEUMATIC PRODUCTS | 5 (3.6%) | 0 (0.0%) | 1 (2.9%) | 4 (4.6%) | |
| ANTITHROMBOTIC AGENTS | 2 (1.4%) | 0 (0.0%) | 0 (0.0%) | 2 (2.3%) | |
| BETA BLOCKING AGENTS | 8 (5.8%) | 0 (0.0%) | 2 (5.7%) | 6 (6.9%) | |
| DIURETICS | 10 (7.2%) | 1 (5.9%) | 2 (5.7%) | 7 (8.0%) | |
| DRUGS FOR ACID RELATED DISORDERS | 4 (2.9%) | 0 (0.0%) | 1 (2.9%) | 3 (3.4%) | |
| DRUGS FOR CONSTIPATION | 2 (1.4%) | 0 (0.0%) | 0 (0.0%) | 2 (2.3%) | |
| DRUGS FOR OBSTRUCTIVE AIRWAY DISEASES | 14 (10.1%) | 2 (11.8%) | 4 (11.4%) | 8 (9.2%) | |
| DRUGS USED IN DIABETES | 9 (6.5%) | 2 (11.8%) | 2 (5.7%) | 5 (5.7%) | |
| OPHTHALMOLOGICALS | 1 (0.7%) | 0 (0.0%) | 0 (0.0%) | 1 (1.1%) | |
| PSYCHOANALEPTICS | 7 (5.0%) | 2 (11.8%) | 1 (2.9%) | 4 (4.6%) | |
| PSYCHOLEPTICS | 2 (1.4%) | 1 (5.9%) | 1 (2.9%) | 0 (0.0%) | |
| UROLOGICALS | 1 (0.7%) | 0 (0.0%) | 1 (2.9%) | 0 (0.0%) | |
| VASOPROTECTIVES | 3 (2.2%) | 0 (0.0%) | 1 (2.9%) | 2 (2.3%) | |

[1]*Chi-square test or Fisher's exact test (f)*

| Table S2. First symptomatology associated with SARS-CoV-2 infection by absence/presence[a] | | | | |
|---|---|---|---|---|
| | Total sample<br>n (%) | ImmunoFormulation cohort<br>n (%) | Control cohort<br>n (%) | p[1] |
| **1.Fever** | **39 (100.0%)** | **20 (100.0%)** | **19 (100.0%)** | <0.0001(f) |
| Absence | 19 (48.7%) | 17 (85.0%) | 2 (10.5%) | |
| Presence | 20 (51.3%) | 3 (15.0%) | 17 (89.5%) | |
| **2.Dry cough** | **39 (100.0%)** | **20 (100.0%)** | **19 (100.0%)** | 0.0011(f) |
| Absence | 23 (59.0%) | 17 (85.0%) | 6 (31.6%) | |
| Presence | 16 (41.0%) | 3 (15.0%) | 13 (68.4%) | |
| **3.Dyspnea** | **39 (100.0%)** | **20 (100.0%)** | **19 (100.0%)** | 0.0004(f) |
| Absence | 27 (69.2%) | 19 (95.0%) | 8 (42.1%) | |
| Presence | 12 (30.8%) | 1 (5.0%) | 11 (57.9%) | |
| **4.Loss of taste and smell** | **39 (100.0%)** | **20 (100.0%)** | **19 (100.0%)** | 0.6050(f) |
| Absence | 35 (89.7%) | 17 (85.0%) | 18 (94.7%) | |
| Presence | 4 (10.3%) | 3 (15.0%) | 1 (5.3%) | |
| **5.Headache** | **39 (100.0%)** | **20 (100.0%)** | **19 (100.0%)** | 0.0915(f) |
| Absence | 32 (82.1%) | 14 (70.0%) | 18 (94.7%) | |
| Presence | 7 (17.9%) | 6 (30.0%) | 1 (5.3%) | |
| **6.Diarrehea** | **39 (100.0%)** | **20 (100.0%)** | **19 (100.0%)** | 1.0000(f) |
| Absence | 35 (89.7%) | 18 (90.0%) | 17 (89.5%) | |
| Presence | 4 (10.3%) | 2 (10.0%) | 2 (10.5%) | |
| **7.Abdominal pain** | **39 (100.0%)** | **20 (100.0%)** | **19 (100.0%)** | 0.1060(f) |
| Absence | 35 (89.7%) | 16 (80.0%) | 19 (100.0%) | |
| Presence | 4 (10.3%) | 4 (20.0%) | 0 (0.0%) | |
| **8.Dermatological findings** | **39 (100.0%)** | **20 (100.0%)** | **19 (100.0%)** | 1.0000(f) |
| Absence | 38 (97.4%) | 19 (95.0%) | 19 (100.0%) | |
| Presence | 1 (2.6%) | 1 (5.0%) | 0 (0.0%) | |
| **9.1. General discomfort[2]** | **39 (100.0%)** | **20 (100.0%)** | **19 (100.0%)** | 0.1060(f) |
| Absence | 35 (89.7%) | 16 (80.0%) | 19 (100.0%) | |
| Presence | 4 (10.3%) | 4 (20.0%) | 0 (0.0%) | |
| **9.2. Throat lesion[2]** | **39 (100.0%)** | **20 (100.0%)** | **19 (100.0%)** | 1.0000(f) |
| Absence | 38 (97.4%) | 19 (95.0%) | 19 (100.0%) | |
| Presence | 1 (2.6%) | 1 (5.0%) | 0 (0.0%) | |
| **9.3. Vomiting[2]** | **39 (100.0%)** | **20 (100.0%)** | **19 (100.0%)** | 1.0000(f) |
| Absence | 38 (97.4%) | 19 (95.0%) | 19 (100.0%) | |
| Presence | 1 (2.6%) | 1 (5.0%) | 0 (0.0%) | |
| **9.4. Weakness[2]** | **39 (100.0%)** | **20 (100.0%)** | **19 (100.0%)** | 0.5273(f) |
| Absence | 18 (46.2%) | 8 (40.0%) | 10 (52.6%) | |
| Presence | 21 (53.8%) | 12 (60.0%) | 9 (47.4%) | |
| **9.5. Sore throat[2]** | **39 (100.0%)** | **20 (100.0%)** | **19 (100.0%)** | 1.0000(f) |
| Absence | 38 (97.4%) | 19 (95.0%) | 19 (100.0%) | |
| Presence | 1 (2.6%) | 1 (5.0%) | 0 (0.0%) | |
| **9.6. Muscular pain[2]** | **39 (100.0%)** | **20 (100.0%)** | **19 (100.0%)** | 1.0000(f) |
| Absence | 37 (94.9%) | 19 (95.0%) | 18 (94.7%) | |

| Table S2. First symptomatology associated with SARS-CoV-2 infection by absence/presence[a] | Total sample n (%) | ImmunoFormulation cohort n (%) | Control cohort n (%) | p[1] |
|---|---|---|---|---|
| Presence | 2 (5.1%) | 1 (5.0%) | 1 (5.3%) | |
| **9.7. Dehydration[2]** | **39 (100.0%)** | **20 (100.0%)** | **19 (100.0%)** | 0.4872(f) |
| Absence | 38 (97.4%) | 20 (100.0%) | 18 (94.7%) | |
| Presence | 1 (2.6%) | 0 (0.0%) | 1 (5.3%) | |
| **9.8. Emesis[2]** | **39 (100.0%)** | **20 (100.0%)** | **19 (100.0%)** | 0.2308(f) |
| Absence | 37 (94.9%) | 20 (100.0%) | 17 (89.5%) | |
| Presence | 2 (5.1%) | 0 (0.0%) | 2 (10.5%) | |
| **9.9. Hypoxemia[2]** | **39 (100.0%)** | **20 (100.0%)** | **19 (100.0%)** | 0.0471(f) |
| Absence | 35 (89.7%) | 20 (100.0%) | 15 (78.9%) | |
| Presence | 4 (10.3%) | 0 (0.0%) | 4 (21.1%) | |
| **9.10. Dysuria[2]** | **39 (100.0%)** | **20 (100.0%)** | **19 (100.0%)** | 0.1060(f) |
| Absence | 36 (92.3%) | 20 (100.0%) | 16 (84.2%) | |
| Presence | 3 (7.7%) | 0 (0.0%) | 3 (15.8%) | |
| **9.11. Pollakiuria[2]** | **39 (100.0%)** | **20 (100.0%)** | **19 (100.0%)** | 0.2308(f) |
| Absence | 37 (94.9%) | 20 (100.0%) | 17 (89.5%) | |
| Presence | 2 (5.1%) | 0 (0.0%) | 2 (10.5%) | |
| **9.12. Sleepiness[2]** | **39 (100.0%)** | **20 (100.0%)** | **19 (100.0%)** | 0.4872(f) |
| Absence | 38 (97.4%) | 20 (100.0%) | 18 (94.7%) | |
| Presence | 1 (2.6%) | 0 (0.0%) | 1 (5.3%) | |
| **9.14. Apathy[2]** | **39 (100.0%)** | **20 (100.0%)** | **19 (100.0%)** | 0.4872(f) |
| Absence | 38 (97.4%) | 20 (100.0%) | 18 (94.7%) | |
| Presence | 1 (2.6%) | 0 (0.0%) | 1 (5.3%) | |
| **9.14. Disorientation[2]** | **39 (100.0%)** | **20 (100.0%)** | **19 (100.0%)** | 0.4872(f) |
| Absence | 38 (97.4%) | 20 (100.0%) | 18 (94.7%) | |
| Presence | 1 (2.6%) | 0 (0.0%) | 1 (5.3%) | |
| **9.15. Anorexia[2]** | **39 (100.0%)** | **20 (100.0%)** | **19 (100.0%)** | 0.2308(f) |
| Absence | 37 (94.9%) | 20 (100.0%) | 17 (89.5%) | |
| Presence | 2 (5.1%) | 0 (0.0%) | 2 (10.5%) | |
| **9.16. Myalgia[2]** | **39 (100.0%)** | **20 (100.0%)** | **19 (100.0%)** | 0.4872(f) |
| Absence | 38 (97.4%) | 20 (100.0%) | 18 (94.7%) | |
| Presence | 1 (2.6%) | 0 (0.0%) | 1 (5.3%) | |
| **9.17. Nasal congestion[2]** | **39 (100.0%)** | **20 (100.0%)** | **19 (100.0%)** | 0.4872(f) |
| Absence | 38 (97.4%) | 20 (100.0%) | 18 (94.7%) | |
| Presence | 1 (2.6%) | 0 (0.0%) | 1 (5.3%) | |

[1] *Fisher exact test (f)*
[2] *Other symptoms: According to MedDRA 23.0 (LLT)*

| Table S3. First symptomatology associated with SARS-CoV-2 infection by severity | | | | |
|---|---|---|---|---|
| | Total sample | ImmunoFormulation cohort | Control cohort | p[1] |
| **1.Fever** | **39 (100.0%)** | **20 (100.0%)** | **19 (100.0%)** | <0.0001(f) |
| Absent | 19 (48.7%) | 17 (85.0%) | 2 (10.5%) | |
| Mild | 10 (25.6%) | 1 (5.0%) | 9 (47.4%) | |
| Moderate | 3 (7.7%) | 0 (0.0%) | 3 (15.8%) | |
| Severe | 7 (17.9%) | 2 (10.0%) | 5 (26.3%) | |
| **2.Dry cough** | **39 (100.0%)** | **20 (100.0%)** | **19 (100.0%)** | 0.0026(f) |
| Absent | 23 (59.0%) | 17 (85.0%) | 6 (31.6%) | |
| Mild | 4 (10.3%) | 1 (5.0%) | 3 (15.8%) | |
| Moderate | 5 (12.8%) | 0 (0.0%) | 5 (26.3%) | |
| Severe | 7 (17.9%) | 2 (10.0%) | 5 (26.3%) | |
| **3.Dyspnea** | **39 (100.0%)** | **20 (100.0%)** | **19 (100.0%)** | 0.0004(f) |
| Absent | 27 (69.2%) | 19 (95.0%) | 8 (42.1%) | |
| Mild | 4 (10.3%) | 0 (0.0%) | 4 (21.1%) | |
| Moderate | 4 (10.3%) | 0 (0.0%) | 4 (21.1%) | |
| Severe | 4 (10.3%) | 1 (5.0%) | 3 (15.7%) | |
| **4.Loss of taste and smell** | **39 (100.0%)** | **20 (100.0%)** | **19 (100.0%)** | 1.0000(f) |
| Absent | 35 (89.7%) | 17 (85.0%) | 18 (94.7%) | |
| Mild | 0 (0.0%) | 0 (0.0%) | 0 (0.0%) | |
| Moderate | 1 (2.6%) | 1 (5.0%) | 0 (0.0%) | |
| Severe | 3 (7.7%) | 2 (10.0%) | 1 (5.3%) | |
| **5.Headache** | **39 (100.0%)** | **20 (100.0%)** | **19 (100.0%)** | 0.1065(f) |
| Absent | 32 (82.1%) | 14 (70.0%) | 18 (94.7%) | |
| Mild | 5 (12.8%) | 4 (20.0%) | 1 (5.3%) | |
| Moderate | 2 (5.1%) | 2 (10.0%) | 0 (0.0%) | |
| Severe | 0 (0.0%) | 0 (0.0%) | 0 (0.0%) | |
| **6.Diarrehea** | **39 (100.0%)** | **20 (100.0%)** | **19 (100.0%)** | 1.0000(f) |
| Absent | 35 (89.7%) | 18 (90.0%) | 17 (89.5%) | |
| Mild | 3 (7.7%) | 1 (5.0%) | 2 (10.5%) | |
| Moderate | 0 (0.0%) | 0 (0.0%) | 0 (0.0%) | |
| Severe | 1 (2.6%) | 1 (5.0%) | 0 (0.0%) | |
| **7.Abdominal pain** | **39 (100.0%)** | **20 (100.0%)** | **19 (100.0%)** | 0.1649(f) |
| Absent | 35 (89.7%) | 16 (80.0%) | 19 (100.0%) | |
| Mild | 3 (7.7%) | 3 (15.0%) | 0 (0.0%) | |
| Moderate | 1 (2.6%) | 1 (5.0%) | 0 (0.0%) | |
| Severe | 0 (0.0%) | 0 (0.0%) | 0 (0.0%) | |
| **8.Dermatological finding** | **39 (100.0%)** | **20 (100.0%)** | **19 (100.0%)** | 1.0000(f) |
| Absent | 38 (97.4%) | 19 (95.0%) | 19 (100.0%) | |
| Mild | 0 (0.0%) | 0 (0.0%) | 0 (0.0%) | |
| Moderate | 0 (0.0%) | 0 (0.0%) | 0 (0.0%) | |
| Severe | 1 (2.6%) | 1 (5.0%) | 0 (0.0%) | |
| **9.1. General discomfort[2]** | **39 (100.0%)** | **20 (100.0%)** | **19 (100.0%)** | 0.2238(f) |
| Absent | 35 (89.7%) | 16 (80.0%) | 19 (100.0%) | |

| Table S3. First symptomatology associated with SARS-CoV-2 infection by severity | | | | |
|---|---|---|---|---|
| | Total sample | ImmunoFormulation cohort | Control cohort | p[1] |
| Mild | 1 (2.6%) | 1 (5.0%) | 0 (0.0%) | |
| Moderate | 1 (2.6%) | 1 (5.0%) | 0 (0.0%) | |
| Severe | 2 (5.1%) | 2 (10.0%) | 0 (0.0%) | |
| **9.2. Throat lesion[2]** | **39 (100.0%)** | **20 (100.0%)** | **19 (100.0%)** | 1.0000(f) |
| Absent | 38 (97.4%) | 19 (95.0%) | 19 (100.0%) | |
| Mild | 0 (0.0%) | 0 (0.0%) | 0 (0.0%) | |
| Moderate | 0 (0.0%) | 0 (0.0%) | 0 (0.0%) | |
| Severe | 1 (2.6%) | 1 (5.0%) | 0 (0.0%) | |
| **9.3. Vomiting[2]** | **39 (100.0%)** | **20 (100.0%)** | **19 (100.0%)** | 1.0000(f) |
| Absent | 38 (97.4%) | 19 (95.0%) | 19 (100.0%) | |
| Mild | 0 (0.0%) | 0 (0.0%) | 0 (0.0%) | |
| Moderate | 0 (0.0%) | 0 (0.0%) | 0 (0.0%) | |
| Severe | 1 (2.6%) | 1 (5.0%) | 0 (0.0%) | |
| **9.4. Weakness[2]** | **39 (100.0%)** | **20 (100.0%)** | **19 (100.0%)** | 0.2063(f) |
| Absent | 18 (46.2%) | 8 (40.0%) | 10 (52.6%) | |
| Mild | 12 (30.8%) | 8 (40.0%) | 4 (21.1%) | |
| Moderate | 2 (5.1%) | 2 (10.0%) | 0 (0.0%) | |
| Severe | 7 (17.9%) | 2 (10.0%) | 5 (26.3%) | |
| **9.5. Sore throat[2]** | **39 (100.0%)** | **20 (100.0%)** | **19 (100.0%)** | 1.0000(f) |
| Absent | 38 (97.4%) | 19 (95.0%) | 19 (100.0%) | |
| Mild | 1 (2.6%) | 1 (5.0%) | 0 (0.0%) | |
| Moderate | 0 (0.0%) | 0 (0.0%) | 0 (0.0%) | |
| Severe | 0 (0.0%) | 0 (0.0%) | 0 (0.0%) | |
| **9.6. Muscular pain[2]** | **39 (100.0%)** | **20 (100.0%)** | **19 (100.0%)** | 1.0000(f) |
| Absent | 37 (94.9%) | 19 (95.0%) | 18 (94.7%) | |
| Mild | 0 (0.0%) | 0 (0.0%) | 0 (0.0%) | |
| Moderate | 1 (2.6%) | 1 (5.0%) | 0 (0.0%) | |
| Severe | 1 (2.6%) | 0 (0.0%) | 1 (5.3%) | |
| **9.7. Dehydration[2]** | **39 (100.0%)** | **20 (100.0%)** | **19 (100.0%)** | 0.4872(f) |
| Absent | 38 (97.4%) | 20 (100.0%) | 18 (94.7%) | |
| Mild | 0 (0.0%) | 0 (0.0%) | 0 (0.0%) | |
| Moderate | 0 (0.0%) | 0 (0.0%) | 0 (0.0%) | |
| Severe | 1 (2.6%) | 0 (0.0%) | 1 (5.3%) | |
| **9.8. Emesis[2]** | **39 (100.0%)** | **20 (100.0%)** | **19 (100.0%)** | 0.2308(f) |
| Absent | 37 (94.9%) | 20 (100.0%) | 17 (89.5%) | |
| Mild | 1 (2.6%) | 0 (0.0%) | 1 (5.3%) | |
| Moderate | 1 (2.6%) | 0 (0.0%) | 1 (5.3%) | |
| Severe | 0 (0.0%) | 0 (0.0%) | 0 (0.0%) | |
| **9.9. Hypoxemia[2]** | **39 (100.0%)** | **20 (100.0%)** | **19 (100.0%)** | 0.0471(f) |
| Absent | 35 (89.7%) | 20 (100.0%) | 15 (78.9%) | |
| Mild | 0 (0.0%) | 0 (0.0%) | 0 (0.0%) | |
| Moderate | 4 (10.3%) | 0 (0.0%) | 4 (21.1%) | |
| Severe | 0 (0.0%) | 0 (0.0%) | 0 (0.0%) | |

| Table S3. First symptomatology associated with SARS-CoV-2 infection by severity | | | | |
|---|---|---|---|---|
| | Total sample | ImmunoFormulation cohort | Control cohort | p[1] |
| **9.10. Dysuria[2]** | **39 (100.0%)** | **20 (100.0%)** | **19 (100.0%)** | 0.1060(f) |
| Absent | 36 (92.3%) | 20 (100.0%) | 16 (84.2%) | |
| Mild | 2 (5.1%) | 0 (0.0%) | 2 (10.5%) | |
| Moderate | 1 (2.6%) | 0 (0.0%) | 1 (5.3%) | |
| Severe | 0 (0.0%) | 0 (0.0%) | 0 (0.0%) | |
| **9.11. Pollakiuria[2]** | **39 (100.0%)** | **20 (100.0%)** | **19 (100.0%)** | 0.2308(f) |
| Absent | 37 (94.9%) | 20 (100.0%) | 17 (89.5%) | |
| Mild | 1 (2.6%) | 0 (0.0%) | 1 (5.3%) | |
| Moderate | 1 (2.6%) | 0 (0.0%) | 1 (5.3%) | |
| Severe | 0 (0.0%) | 0 (0.0%) | 0 (0.0%) | |
| **9.12. Sleepiness[2]** | **39 (100.0%)** | **20 (100.0%)** | **19 (100.0%)** | 0.4872(f) |
| Absent | 38 (97.4%) | 20 (100.0%) | 18 (94.7%) | |
| Mild | 0 (0.0%) | 0 (0.0%) | 0 (0.0%) | |
| Moderate | 1 (2.6%) | 0 (0.0%) | 1 (5.3%) | |
| Severe | 0 (0.0%) | 0 (0.0%) | 0 (0.0%) | |
| **9.13. Apathy[2]** | **39 (100.0%)** | **20 (100.0%)** | **19 (100.0%)** | 0.4872(f) |
| Absent | 38 (97.4%) | 20 (100.0%) | 18 (94.7%) | |
| Mild | 0 (0.0%) | 0 (0.0%) | 0 (0.0%) | |
| Moderate | 0 (0.0%) | 0 (0.0%) | 0 (0.0%) | |
| Severe | 1 (2.6%) | 0 (0.0%) | 1 (5.3%) | |
| **9.14. Disorientation[2]** | **39 (100.0%)** | **20 (100.0%)** | **19 (100.0%)** | 0.4872(f) |
| Absent | 38 (97.4%) | 20 (100.0%) | 18 (94.7%) | |
| Mild | 1 (2.6%) | 0 (0.0%) | 1 (5.3%) | |
| Moderate | 0 (0.0%) | 0 (0.0%) | 0 (0.0%) | |
| Severe | 0 (0.0%) | 0 (0.0%) | 0 (0.0%) | |
| **9.15. Anorexia[2]** | **39 (100.0%)** | **20 (100.0%)** | **19 (100.0%)** | 0.2308(f) |
| Absent | 37 (94.9%) | 20 (100.0%) | 17 (89.5%) | |
| Mild | 0 (0.0%) | 0 (0.0%) | 0 (0.0%) | |
| Moderate | 0 (0.0%) | 0 (0.0%) | 0 (0.0%) | |
| Severe | 2 (5.1%) | 0 (0.0%) | 2 (10.5%) | |
| **9.16. Myalgia[2]** | **39 (100.0%)** | **20 (100.0%)** | **19 (100.0%)** | 0.4872(f) |
| Absent | 38 (97.4%) | 20 (100.0%) | 18 (94.7%) | |
| Mild | 0 (0.0%) | 0 (0.0%) | 0 (0.0%) | |
| Moderate | 0 (0.0%) | 0 (0.0%) | 0 (0.0%) | |
| Severe | 1 (2.6%) | 0 (0.0%) | 1 (5.3%) | |
| **9.17. Nasal congestion[2]** | **39 (100.0%)** | **20 (100.0%)** | **19 (100.0%)** | 0.4872(f) |
| Absent | 38 (97.4%) | 20 (100.0%) | 18 (94.7%) | |
| Mild | 1 (2.6%) | 0 (0.0%) | 1 (5.3%) | |
| Moderate | 0 (0.0%) | 0 (0.0%) | 0 (0.0%) | |
| Severe | 0 (0.0%) | 0 (0.0%) | 0 (0.0%) | |

[1] *Fisher exact test (f)*

| Table S4. Recovery duration of each symptom associated with COVID-19 by symptoms | | | | |
|---|---|---|---|---|
| | Total sample | ImmunoFormulation cohort | Control cohort | p[1] |
| **TOTAL RECOVERY FROM START OF THE FIRST SYMPTOM[a]** | | | | |
| **1. Fever** | | | | |
| **Days with some symptoms to the end of the observational period** | | | | 0 |
| Mean (SD) | 12.08 (10.90) | 3.35 (2.87) | 21.78 (7.75) | <0.0001 |
| 95%CI | (8.50 ; 15.66) | (2.01 ; 4.69) | (17.92 ; 25.63) | |
| Median (P25 ; P75) | 10.00 (2.00 ; 24.00) | 2.00 (2.00 ; 3.00) | 24.00 (14.00 ; 30.00) | |
| (Min ; Max) | (1.00 ; 31.00) | (1.00 ; 13.00) | (11.00 ; 31.00) | |
| N valid | 38 | 20 | 18 | |
| **PATIENTS RECOVERED TO THE END OF THE OBSERVATIONAL PERIOD,** n (%) | **38 (100.0%)** | **20 (100.0%)** | **18 (100.0%)** | 0.0067(f) |
| Yes | 32 (84.2%) | 20 (100.0%) | 12 (66.7%) | |
| No | 6 (15.8%) | 0 (0.0%) | 6 (33.3%) | |
| **PATIENT RECOVERED- Days with some symptoms to the end of the observational period** | | | | |
| Mean (SD) | 9.22 (8.96) | 3.35 (2.87) | 19.00 (6.71) | <0.0001 |
| 95%CI | (5.99 ; 12.45) | (2.01 ; 4.69) | (14.73 ; 23.27) | |
| Median (P25 ; P75) | 3.00 (2.00 ; 14.00) | 2.00 (2.00 ; 3.00) | 18.50 (13.00 ; 24.50) | |
| (Min ; Max) | (1.00 ; 30.00) | (1.00 ; 13.00) | (11.00 ; 30.00) | |
| N valid | 32 | 20 | 12 | |
| **2. Dry Cough** | | | | |
| **Days with some symptoms to the end of the observational period** | | | | |
| Mean (SD) | 15.71 (11.37) | 6.15 (6.52) | 24.00 (7.39) | <0.0001 |
| 95%CI | (11.30 ; 20.12) | (2.22 ; 10.09) | (19.91 ; 28.09) | |
| Median (P25 ; P75) | 16.50 (4.00 ; 27.50) | 4.00 (2.00 ; 6.00) | 25.00 (20.00 ; 30.00) | |
| (Min ; Max) | (2.00 ; 30.00) | (2.00 ; 25.00) | (6.00 ; 30.00) | |
| N valid | 28 | 13 | 15 | |
| **PATIENTS RECOVERED TO THE END OF THE OBSERVATIONAL PERIOD,** n (%) | **28 (100.0%)** | **13 (100.0%)** | **15 (100.0%)** | 0.0069(f) |
| Yes | 21 (75.0%) | 13 (100.0%) | 8 (53.3%) | |
| No | 7 (25.0%) | 0 (0.0%) | 7 (46.7%) | |
| **PATIENT RECOVERED- Days with some symptoms to the end of the observational period** | | | | |
| Mean (SD) | 11.29 (9.49) | 6.15 (6.52) | 19.63 (7.50) | 0.0025 |
| 95%CI | (6.96 ; 15.61) | (2.22 ; 10.09) | (13.35 ; 25.90) | |
| Median (P25 ; P75) | 6.00 (3.00 ; 20.00) | 4.00 (2.00 ; 6.00) | 20.50 (16.00 ; 24.00) | |
| (Min ; Max) | (2.00 ; 30.00) | (2.00 ; 25.00) | (6.00 ; 30.00) | |
| N valid | 21 | 13 | 8 | |
| **3. Dyspnea** | | | | |
| **Days with some symptoms to the end of the observational period** | | | | |
| Mean (SD) | 17.47 (8.94) | 5.67 (6.35) | 20.00 (7.29) | 0.0315 |
| 95%CI | (12.88 ; 22.07) | (0.00 ; 21.44) | (15.79 ; 24.21) | |
| Median (P25 ; P75) | 20.00 (12.00 ; 24.00) | 2.00 (2.00 ; 13.00) | 20.50 (13.00 ; 25.00) | |

| Table S4. Recovery duration of each symptom associated with COVID-19 by symptoms | | | | |
|---|---|---|---|---|
| | Total sample | ImmunoFormulation cohort | Control cohort | p[1] |
| (Min ; Max)<br>N valid | (2.00 ; 30.00)<br>17 | (2.00 ; 13.00)<br>3 | (7.00 ; 30.00)<br>14 | |
| **PATIENTS RECOVERED TO THE END OF THE OBSERVATIONAL PERIOD,** n (%) | **17 (100.0%)** | **3 (100.0%)** | **14 (100.0%)** | 0.5412(f) |
| Yes | 13 (76.5%) | 3 (100.0%) | 10 (71.4%) | |
| No | 4 (23.5%) | 0 (0.0%) | 4 (28.6%) | |
| **PATIENT RECOVERED-Days with some symptoms to the end of the observational period** | | | | |
| Mean (SD) | 17.31 (9.23) | 5.67 (6.35) | 20.80 (6.78) | 0.0341 |
| 95%CI | (11.73 ; 22.89) | (0.00 ; 21.44) | (15.95 ; 25.65) | |
| Median (P25 ; P75) | 20.00 (12.00 ; 24.00) | 2.00 (2.00 ; 13.00) | 22.00 (20.00 ; 25.00) | |
| (Min ; Max) | (2.00 ; 30.00) | (2.00 ; 13.00) | (7.00 ; 30.00) | |
| N valid | 13 | 3 | 10 | |
| **4. Loss of taste and smell** | | | | |
| **Days with some symptoms to the end of the observational period** | | | | |
| Mean (SD) | 22.31 (7.04) | 21.55 (7.27) | 26.50 (4.95) | 0.4860 |
| 95%CI | (18.05 ; 26.56) | (16.66 ; 26.43) | (0.00 ; 70.97) | |
| Median (P25 ; P75) | 23.00 (15.00 ; 29.00) | 23.00 (14.00 ; 29.00) | 26.50 (23.00 ; 30.00) | |
| (Min ; Max) | (11.00 ; 31.00) | (11.00 ; 31.00) | (23.00 ; 30.00) | |
| N valid | 13 | 11 | 2 | |
| **PATIENTS RECOVERED TO THE END OF THE OBSERVATIONAL PERIOD,** n (%) | **13 (100.0%)** | **11 (100.0%)** | **2 (100.0%)** | 0.0385(f) |
| Yes | 10 (76.9%) | 10 (90.9%) | 0 (0.0%) | |
| No | 3 (23.1%) | 1 (9.1%) | 2 (100.0%) | |
| **PATIENT RECOVERED-Days with some symptoms to the end of the observational period** | | | | |
| Mean (SD) | 20.60 (6.92) | 20.60 (6.92) | -- | -- |
| 95%CI | (15.65 ; 25.55) | (15.65 ; 25.55) | | |
| Median (P25 ; P75) | 21.50 (14.00 ; 26.00) | 21.50 (14.00 ; 26.00) | | |
| (Min ; Max) | (11.00 ; 31.00) | (11.00 ; 31.00) | | |
| N valid | 10 | 10 | | |
| **5. Headache** | | | | |
| **Days with some symptoms to the end of the observational period** | | | | |
| Mean (SD) | 10.30 (8.87) | 6.25 (1.98) | 26.50 (4.95) | 0.0467 |
| 95%CI | (3.95 ; 16.65) | (4.59 ; 7.91) | (0.00 ; 70.97) | |
| Median (P25 ; P75) | 7.00 (6.00 ; 8.00) | 7.00 (5.50 ; 7.50) | 26.50 (23.00 ; 30.00) | |
| (Min ; Max) | (2.00 ; 30.00) | (2.00 ; 8.00) | (23.00 ; 30.00) | |
| N valid | 10 | 8 | 2 | |
| **PATIENTS RECOVERED TO THE END OF THE OBSERVATIONAL PERIOD,** n (%) | **10 (100.0%)** | **8 (100.0%)** | **2 (100.0%)** | 0.0222(f) |
| Yes | 8 (80.0%) | 8 (100.0%) | 0 (0.0%) | |
| No | 2 (20.0%) | 0 (0.0%) | 2 (100.0%) | |
| **PATIENT RECOVERED-Days with some symptoms to the end of the observational period** | | | | |

| Table S4. Recovery duration of each symptom associated with COVID-19 by symptoms | | | | |
|---|---|---|---|---|
| | **Total sample** | **ImmunoFormulation cohort** | **Control cohort** | p[1] |
| Mean (SD) | 6.25 (1.98) | 6.25 (1.98) | -- | -- |
| 95%CI | (4.59 ; 7.91) | (4.59 ; 7.91) | | |
| Median (P25 ; P75) | 7.00 (5.50 ; 7.50) | 7.00 (5.50 ; 7.50) | | |
| (Min ; Max) | (2.00 ; 8.00) | (2.00 ; 8.00) | | |
| N valid | 8 | 8 | | |
| **6. Diarrhea** | | | | |
| **Days with some symptoms to the end of the observational period** | | | | |
| Mean (SD) | 17.00 (9.50) | 8.75 (4.35) | 25.25 (3.20) | 0.0294 |
| 95%CI | (9.06 ; 24.94) | (1.83 ; 15.67) | (20.16 ; 30.34) | |
| Median (P25 ; P75) | 18.00 (9.50 ; 24.00) | 9.50 (5.50 ; 12.00) | 24.00 (23.50 ; 27.00) | |
| (Min ; Max) | (3.00 ; 30.00) | (3.00 ; 13.00) | (23.00 ; 30.00) | |
| N valid | 8 | 4 | 4 | |
| **PATIENTS RECOVERED TO THE END OF THE OBSERVATIONAL PERIOD,** n (%) | **8 (100.0%)** | **4 (100.0%)** | **4 (100.0%)** | 0.1429(f) |
| Yes | 5 (62.5%) | 4 (100.0%) | 1 (25.0%) | |
| No | 3 (37.5%) | 0 (0.0%) | 3 (75.0%) | |
| **PATIENT RECOVERED- Days with some symptoms to the end of the observational period** | | | | |
| Mean (SD) | 11.80 (7.79) | 8.75 (4.35) | 24.00 (.) | 0.2888 |
| 95%CI | (2.13 ; 21.47) | (1.83 ; 15.67) | (. ; .) | |
| Median (P25 ; P75) | 11.00 (8.00 ; 13.00) | 9.50 (5.50 ; 12.00) | 24.00 (24.00 ; 24.00) | |
| (Min ; Max) | (3.00 ; 24.00) | (3.00 ; 13.00) | (24.00 ; 24.00) | |
| N valid | 5 | 4 | 1 | |
| **7. Abdominal pain** | | | | |
| **Days with some symptoms to the end of the observational period** | | | | |
| Mean (SD) | 6.80 (2.17) | 6.80 (2.17) | . (.) | -- |
| 95%CI | (4.11 ; 9.49) | (4.11 ; 9.49) | (. ; .) | |
| Median (P25 ; P75) | 8.00 (7.00 ; 8.00) | 8.00 (7.00 ; 8.00) | . (. ; .) | |
| (Min ; Max) | (3.00 ; 8.00) | (3.00 ; 8.00) | (. ; .) | |
| N valid | 5 | 5 | 0 | |
| **PATIENTS RECOVERED TO THE END OF THE OBSERVATIONAL PERIOD,** n (%) | **5 (100.0%)** | **5 (100.0%)** | **--** | **--** |
| Yes | 5 (100.0%) | 5 (100.0%) | | |
| No | 5 (100.0%) | 5 (100.0%) | | |
| **PATIENT RECOVERED- Days with some symptoms to the end of the observational period** | | | | |
| Mean (SD) | 6.80 (2.17) | 6.80 (2.17) | -- | -- |
| 95%CI | (4.11 ; 9.49) | (4.11 ; 9.49) | | |
| Median (P25 ; P75) | 8.00 (7.00 ; 8.00) | 8.00 (7.00 ; 8.00) | | |
| (Min ; Max) | (3.00 ; 8.00) | (3.00 ; 8.00) | | |
| N valid | 5 | 5 | | |
| **8. Dermatological findings** | | | | |
| **Days with some symptoms to the end of the observational period** | | | | |
| Mean (SD) | 31.00 (.) | 31.00 (.) | . (.) | -- |
| 95%CI | (. ; .) | (. ; .) | (. ; .) | |
| Median (P25 ; P75) | 31.00 (31.00 ; 31.00) | 31.00 (31.00 ; 31.00) | . (. ; .) | |

| Table S4. Recovery duration of each symptom associated with COVID-19 by symptoms | | | | |
|---|---|---|---|---|
| | **Total sample** | **ImmunoFormulation cohort** | **Control cohort** | p[1] |
| (Min ; Max)<br>N valid | (31.00 ; 31.00)<br>1 | (31.00 ; 31.00)<br>1 | (. ; .)<br>0 | |
| **PATIENTS RECOVERED TO THE END OF THE OBSERVATIONAL PERIOD,** n (%)<br>Yes<br>No | **1 (100.0%)**<br>0 (0.0%)<br>1 (100.0%) | **1 (100.0%)**<br>0 (0.0%)<br>1 (100.0%) | **--** | **--** |
| **PATIENT RECOVERED-Days with some symptoms to the end of the observational period**<br>Mean (SD)<br>95%CI<br>Median (P25 ; P75)<br>(Min ; Max)<br>N valid | -- | -- | -- | -- |
| **9.1. General discomfort[3]** | | | | |
| **Days with some symptoms to the end of the observational period**<br>Mean (SD)<br>95%CI<br>Median (P25 ; P75)<br>(Min ; Max)<br>N valid | 15.75 (10.37)<br>(0.00 ; 32.25)<br>12.00 (9.50 ; 22.00)<br>(8.00 ; 31.00)<br>4 | 15.75 (10.37)<br>(0.00 ; 32.25)<br>12.00 (9.50 ; 22.00)<br>(8.00 ; 31.00)<br>4 | . (.)<br>(. ; .)<br>. (. ; .)<br>(. ; .)<br>0 | -- |
| **PATIENTS RECOVERED TO THE END OF THE OBSERVATIONAL PERIOD,** n (%)<br>Yes<br>No | **4 (100.0%)**<br>3 (75.0%)<br>1 (25.0%) | **4 (100.0%)**<br>3 (75.0%)<br>1 (25.0%) | **--** | -- |
| **PATIENT RECOVERED-Days with some symptoms to the end of the observational period**<br>Mean (SD)<br>95%CI<br>Median (P25 ; P75)<br>(Min ; Max)<br>N valid | 10.67 (2.52)<br>(4.42 ; 16.92)<br>11.00 (8.00 ; 13.00)<br>(8.00 ; 13.00)<br>3 | 10.67 (2.52)<br>(4.42 ; 16.92)<br>11.00 (8.00 ; 13.00)<br>(8.00 ; 13.00)<br>3 | -- | -- |
| **9.2. Throat lesion[3]** | | | | |
| **Days with some symptoms to the end of the observational period**<br>Mean (SD)<br>95%CI<br>Median (P25 ; P75)<br>(Min ; Max)<br>N valid | 13.00 (.)<br>(. ; .)<br>13.00 (13.00 ; 13.00)<br>(13.00 ; 13.00)<br>1 | 13.00 (.)<br>(. ; .)<br>13.00 (13.00 ; 13.00)<br>(13.00 ; 13.00)<br>1 | . (.)<br>(. ; .)<br>. (. ; .)<br>(. ; .)<br>0 | -- |
| **PATIENTS RECOVERED TO THE END OF THE OBSERVATIONAL PERIOD,** n (%)<br>Yes<br>No | **1 (100.0%)**<br>1 (100.0%)<br>0 (0.0%) | **1 (100.0%)**<br>1 (100.0%)<br>0 (0.0%) | **--** | **--** |
| **PATIENT RECOVERED-Days with some symptoms to the end of the observational period**<br>Mean (SD)<br>95%CI<br>Median (P25 ; P75) | 13.00 (.)<br>(. ; .)<br>13.00 (13.00 ; 13.00) | 13.00 (.)<br>(. ; .)<br>13.00 (13.00 ; 13.00) | -- | -- |

| Table S4. Recovery duration of each symptom associated with COVID-19 by symptoms | | | | |
|---|---|---|---|---|
| | **Total sample** | **ImmunoFormulation cohort** | **Control cohort** | p[1] |
| (Min ; Max) | (13.00 ; 13.00) | (13.00 ; 13.00) | | |
| N valid | 1 | 1 | | |
| **9.3. Vomiting[3]** | | | | |
| **Days with some symptoms to the end of the observational period** | | | | |
| Mean (SD) | 13.00 (.) | 13.00 (.) | . (.) | -- |
| 95%CI | (. ; .) | (. ; .) | (. ; .) | |
| Median (P25 ; P75) | 13.00 (13.00 ; 13.00) | 13.00 (13.00 ; 13.00) | . (. ; .) | |
| (Min ; Max) | (13.00 ; 13.00) | (13.00 ; 13.00) | (. ; .) | |
| N valid | 1 | 1 | 0 | |
| **PATIENTS RECOVERED TO THE END OF THE OBSERVATIONAL PERIOD,** n (%) | **1 (100.0%)** | **1 (100.0%)** | **--** | -- |
| Yes | 1 (100.0%) | 1 (100.0%) | | |
| No | 0 (0.0%) | 0 (0.0%) | | |
| **PATIENT RECOVERED-Days with some symptoms to the end of the observational period** | | | | |
| Mean (SD) | 13.00 (.) | 13.00 (.) | . (.) | -- |
| 95%CI | (. ; .) | (. ; .) | (. ; .) | |
| Median (P25 ; P75) | 13.00 (13.00 ; 13.00) | 13.00 (13.00 ; 13.00) | . (. ; .) | |
| (Min ; Max) | (13.00 ; 13.00) | (13.00 ; 13.00) | (. ; .) | |
| N valid | 1 | 1 | 0 | |
| **9.4. Weakness[3]** | | | | |
| **Days with some symptoms to the end of the observational period** | | | | |
| Mean (SD) | 14.64 (10.19) | 7.42 (1.08) | 23.30 (9.37) | 0.0060 |
| 95%CI | (10.12 ; 19.15) | (6.73 ; 8.11) | (16.60 ; 30.00) | |
| Median (P25 ; P75) | 8.00 (7.00 ; 26.00) | 7.00 (7.00 ; 8.00) | 27.50 (21.00 ; 30.00) | |
| (Min ; Max) | (6.00 ; 30.00) | (6.00 ; 10.00) | (6.00 ; 30.00) | |
| N valid | 22 | 12 | 10 | |
| **PATIENTS RECOVERED TO THE END OF THE OBSERVATIONAL PERIOD,** n (%) | **22 (100.0%)** | **12 (100.0%)** | **10 (100.0%)** | 0.0096(f) |
| Yes | 17 (77.3%) | 12 (100.0%) | 5 (50.0%) | |
| No | 5 (22.7%) | 0 (0.0%) | 5 (50.0%) | |
| **PATIENT RECOVERED-Days with some symptoms to the end of the observational period** | | | | |
| Mean (SD) | 11.47 (7.93) | 7.42 (1.08) | 21.20 (8.98) | 0.0464 |
| 95%CI | (7.39 ; 15.55) | (6.73 ; 8.11) | (10.05 ; 32.35) | |
| Median (P25 ; P75) | 8.00 (7.00 ; 10.00) | 7.00 (7.00 ; 8.00) | 24.00 (21.00 ; 26.00) | |
| (Min ; Max) | (6.00 ; 29.00) | (6.00 ; 10.00) | (6.00 ; 29.00) | |
| N valid | 17 | 12 | 5 | |
| **9.5. Sore throat[3]** | | | | |
| **Days with some symptoms to the end of the observational period** | | | | |
| Mean (SD) | 7.00 (.) | 7.00 (.) | . (.) | -- |
| 95%CI | (. ; .) | (. ; .) | (. ; .) | |
| Median (P25 ; P75) | 7.00 (7.00 ; 7.00) | 7.00 (7.00 ; 7.00) | . (. ; .) | |
| (Min ; Max) | (7.00 ; 7.00) | (7.00 ; 7.00) | (. ; .) | |
| N valid | 1 | 1 | 0 | |

**Table S4. Recovery duration of each symptom associated with COVID-19 by symptoms**

| | Total sample | ImmunoFormulation cohort | Control cohort | p[1] |
|---|---|---|---|---|
| **PATIENTS RECOVERED TO THE END OF THE OBSERVATIONAL PERIOD,** n (%) | **1 (100.0%)** | **1 (100.0%)** | **--** | **--** |
| Yes | 1 (100.0%) | 1 (100.0%) | | |
| No | 0 (0.0%) | 0 (0.0%) | | |
| **PATIENT RECOVERED- Days with some symptoms to the end of the observational period** | | | | |
| Mean (SD) | 7.00 (.) | 7.00 (.) | . (.) | -- |
| 95%CI | (. ; .) | (. ; .) | (. ; .) | |
| Median (P25 ; P75) | 7.00 (7.00 ; 7.00) | 7.00 (7.00 ; 7.00) | . (. ; .) | |
| (Min ; Max) | (7.00 ; 7.00) | (7.00 ; 7.00) | (. ; .) | |
| N valid | 1 | 1 | 0 | |
| **9.6. Muscular pain[3]** | | | | |
| **Days with some symptoms to the end of the observational period** | | | | |
| Mean (SD) | 10.50 (3.54) | 8.00 (.) | 13.00 (.) | -- |
| 95%CI | (0.00 ; 42.27) | (. ; .) | (. ; .) | |
| Median (P25 ; P75) | 10.50 (8.00 ; 13.00) | 8.00 (8.00 ; 8.00) | 13.00 (13.00 ; 13.00) | |
| (Min ; Max) | (8.00 ; 13.00) | (8.00 ; 8.00) | (13.00 ; 13.00) | |
| N valid | 2 | 1 | 1 | |
| **PATIENTS RECOVERED TO THE END OF THE OBSERVATIONAL PERIOD,** n (%) | **2 (100.0%)** | **1 (100.0%)** | **1 (100.0%)** | -- |
| Yes | 2 (100.0%) | 1 (100.0%) | 1 (100.0%) | |
| No | 0 (0.0%) | 0 (0.0%) | 0 (0.0%) | |
| **PATIENT RECOVERED- Days with some symptoms to the end of the observational period** | | | | |
| Mean (SD) | 10.50 (3.54) | 8.00 (.) | 13.00 (.) | -- |
| 95%CI | (0.00 ; 42.27) | (. ; .) | (. ; .) | |
| Median (P25 ; P75) | 10.50 (8.00 ; 13.00) | 8.00 (8.00 ; 8.00) | 13.00 (13.00 ; 13.00) | |
| (Min ; Max) | (8.00 ; 13.00) | (8.00 ; 8.00) | (13.00 ; 13.00) | |
| N valid | 2 | 1 | 1 | |
| **9.7. Dehydration[3]** | | | | |
| **Days with some symptoms to the end of the observational period** | | | | |
| Mean (SD) | 31.00 (.) | . (.) | 31.00 (.) | -- |
| 95%CI | (. ; .) | (. ; .) | (. ; .) | |
| Median (P25 ; P75) | 31.00 (31.00 ; 31.00) | . (. ; .) | 31.00 (31.00 ; 31.00) | |
| (Min ; Max) | (31.00 ; 31.00) | (. ; .) | (31.00 ; 31.00) | |
| N valid | 1 | 0 | 1 | |
| **PATIENTS RECOVERED TO THE END OF THE OBSERVATIONAL PERIOD,** n (%) | **1 (100.0%)** | **-** | **1 (100.0%)** | -- |
| Yes | 0 (0.0%) | | 0 (0.0%) | |
| No | 1 (100.0%) | | 1 (100.0%) | |
| **PATIENT RECOVERED- Days with some symptoms to the end of the observational period** | | | | |
| Mean (SD) | -- | -- | -- | -- |
| 95%CI | | | | |

| Table S4. Recovery duration of each symptom associated with COVID-19 by symptoms | Total sample | ImmunoFormulation cohort | Control cohort | p[1] |
|---|---|---|---|---|
| Median (P25 ; P75)<br>(Min ; Max)<br>N valid | | | | |
| **9.8. Emesis[3]** | | | | |
| **Days with some symptoms to the end of the observational period** | | | | |
| Mean (SD) | 27.00 (4.24) | . (.) | 27.00 (4.24) | -- |
| 95%CI | (0.00 ; 65.12) | (. ; .) | (0.00 ; 65.12) | |
| Median (P25 ; P75) | 27.00 (24.00 ; 30.00) | . (. ; .) | 27.00 (24.00 ; 30.00) | |
| (Min ; Max) | (24.00 ; 30.00) | (. ; .) | (24.00 ; 30.00) | |
| N valid | 2 | 0 | 2 | |
| **PATIENTS RECOVERED TO THE END OF THE OBSERVATIONAL PERIOD,** n (%) | **2 (100.0%)** | -- | **2 (100.0%)** | -- |
| Yes | 1 (50.0%) | | 1 (50.0%) | |
| No | 1 (50.0%) | | 1 (50.0%) | |
| **PATIENT RECOVERED- Days with some symptoms to the end of the observational period** | | | | |
| Mean (SD) | 24.00 (.) | -- | 24.00 (.) | -- |
| 95%CI | (. ; .) | | (. ; .) | |
| Median (P25 ; P75) | 24.00 (24.00 ; 24.00) | | 24.00 (24.00 ; 24.00) | |
| (Min ; Max) | (24.00 ; 24.00) | | (24.00 ; 24.00) | |
| N valid | 1 | | 1 | |
| **9.9. Hypoxemia[3]** | | | | |
| **Days with some symptoms to the end of the observational period** | | | | |
| Mean (SD) | 22.50 (8.10) | . (.) | 22.50 (8.10) | -- |
| 95%CI | (9.61 ; 35.39) | (. ; .) | (9.61 ; 35.39) | |
| Median (P25 ; P75) | 24.50 (17.50 ; 27.50) | . (. ; .) | 24.50 (17.50 ; 27.50) | |
| (Min ; Max) | (11.00 ; 30.00) | (. ; .) | (11.00 ; 30.00) | |
| N valid | 4 | 0 | 4 | |
| **PATIENTS RECOVERED TO THE END OF THE OBSERVATIONAL PERIOD,** n (%) | **4 (100.0%)** | -- | **4 (100.0%)** | -- |
| Yes | 3 (75.0%) | | 3 (75.0%) | |
| No | 1 (25.0%) | | 1 (25.0%) | |
| **PATIENT RECOVERED- Days with some symptoms to the end of the observational period** | | | | |
| Mean (SD) | 20.00 (7.81) | -- | 20.00 (7.81) | -- |
| 95%CI | (0.60 ; 39.40) | | (0.60 ; 39.40) | |
| Median (P25 ; P75) | 24.00 (11.00 ; 25.00) | | 24.00 (11.00 ; 25.00) | |
| (Min ; Max) | (11.00 ; 25.00) | | (11.00 ; 25.00) | |
| N valid | 3 | | 3 | |
| **9.10. Dysuria[3]** | | | | |
| **Days with some symptoms to the end of the observational period** | | | | |
| Mean (SD) | 25.67 (3.79) | . (.) | 25.67 (3.79) | -- |
| 95%CI | (16.26 ; 35.07) | (. ; .) | (16.26 ; 35.07) | |
| Median (P25 ; P75) | 24.00 (23.00 ; 30.00) | . (. ; .) | 24.00 (23.00 ; 30.00) | |

| Table S4. Recovery duration of each symptom associated with COVID-19 by symptoms | | | | |
|---|---|---|---|---|
| | **Total sample** | **ImmunoFormulation cohort** | **Control cohort** | p[1] |
| (Min ; Max) | (23.00 ; 30.00) | (. ; .) | (23.00 ; 30.00) | |
| N valid | 3 | 0 | 3 | |
| **PATIENTS RECOVERED TO THE END OF THE OBSERVATIONAL PERIOD,** n (%) | **3 (100.0%)** | -- | **3 (100.0%)** | -- |
| Yes | 2 (66.7%) | | 2 (66.7%) | |
| No | 1 (33.3%) | | 1 (33.3%) | |
| **PATIENT RECOVERED-Days with some symptoms to the end of the observational period** | | | | |
| Mean (SD) | 23.50 (0.71) | -- | 23.50 (0.71) | -- |
| 95%CI | (17.15 ; 29.85) | | (17.15 ; 29.85) | |
| Median (P25 ; P75) | 23.50 (23.00 ; 24.00) | | 23.50 (23.00 ; 24.00) | |
| (Min ; Max) | (23.00 ; 24.00) | | (23.00 ; 24.00) | |
| N valid | 2 | | 2 | |
| **9.11. Pollakiuria[3]** | | | | |
| **Days with some symptoms to the end of the observational period** | | | | |
| Mean (SD) | 27.00 (4.24) | . (.) | 27.00 (4.24) | -- |
| 95%CI | (0.00 ; 65.12) | (. ; .) | (0.00 ; 65.12) | |
| Median (P25 ; P75) | 27.00 (24.00 ; 30.00) | . (. ; .) | 27.00 (24.00 ; 30.00) | |
| (Min ; Max) | (24.00 ; 30.00) | (. ; .) | (24.00 ; 30.00) | |
| N valid | 2 | 0 | 2 | |
| **PATIENTS RECOVERED TO THE END OF THE OBSERVATIONAL PERIOD,** n (%) | **2 (100.0%)** | -- | **2 (100.0%)** | -- |
| Yes | 1 (50.0%) | | 1 (50.0%) | |
| No | 1 (50.0%) | | 1 (50.0%) | |
| **PATIENT RECOVERED-Days with some symptoms to the end of the observational period** | | | | |
| Mean (SD) | 24.00 (.) | -- | 24.00 (.) | -- |
| 95%CI | (. ; .) | | (. ; .) | |
| Median (P25 ; P75) | 24.00 (24.00 ; 24.00) | | 24.00 (24.00 ; 24.00) | |
| (Min ; Max) | (24.00 ; 24.00) | | (24.00 ; 24.00) | |
| N valid | 1 | | 1 | |
| **9.12. Sleepiness[3]** | | | | |
| **Days with some symptoms to the end of the observational period** | | | | |
| Mean (SD) | 30.00 (.) | . (.) | 30.00 (.) | -- |
| 95%CI | (. ; .) | (. ; .) | (. ; .) | |
| Median (P25 ; P75) | 30.00 (30.00 ; 30.00) | . (. ; .) | 30.00 (30.00 ; 30.00) | |
| (Min ; Max) | (30.00 ; 30.00) | (. ; .) | (30.00 ; 30.00) | |
| N valid | 1 | 0 | 1 | |
| **PATIENTS RECOVERED TO THE END OF THE OBSERVATIONAL PERIOD,** n (%) | **1 (100.0%)** | **--** | **1 (100.0%)** | -- |
| Yes | 0 (0.0%) | | 0 (0.0%) | |
| No | 1 (100.0%) | | 1 (100.0%) | |

| Table S4. Recovery duration of each symptom associated with COVID-19 by symptoms | | | | |
|---|---|---|---|---|
| | **Total sample** | **ImmunoFormulation cohort** | **Control cohort** | p[1] |
| **PATIENT RECOVERED- Days with some symptoms to the end of the observational period**<br>Mean (SD)<br>95%CI<br>Median (P25 ; P75)<br>(Min ; Max)<br>N valid | -- | -- | -- | -- |
| **9.13. Apathy[3]** | | | | |
| **Days with some symptoms to the end of the observational period**<br>Mean (SD)<br>95%CI<br>Median (P25 ; P75)<br>(Min ; Max)<br>N valid | 24.00 (.)<br>(. ; .)<br>24.00 (24.00 ; 24.00)<br>(24.00 ; 24.00)<br>1 | . (.)<br>(. ; .)<br>. (. ; .)<br>(. ; .)<br>0 | 24.00 (.)<br>(. ; .)<br>24.00 (24.00 ; 24.00)<br>(24.00 ; 24.00)<br>1 | -- |
| **PATIENTS RECOVERED TO THE END OF THE OBSERVATIONAL PERIOD,** n (%)<br>Yes<br>No | **1 (100.0%)**<br>1 (100.0%)<br>0 (0.0%) | -- | **1 (100.0%)**<br>1 (100.0%)<br>0 (0.0%) | -- |
| **PATIENT RECOVERED- Days with some symptoms to the end of the observational period**<br>Mean (SD)<br>95%CI<br>Median (P25 ; P75)<br>(Min ; Max)<br>N valid | 24.00 (.)<br>(. ; .)<br>24.00 (24.00 ; 24.00)<br>(24.00 ; 24.00)<br>1 | -- | 24.00 (.)<br>(. ; .)<br>24.00 (24.00 ; 24.00)<br>(24.00 ; 24.00)<br>1 | -- |
| **9.14. Disorientation[3]** | | | | |
| **Days with some symptoms to the end of the observational period**<br>Mean (SD)<br>95%CI<br>Median (P25 ; P75)<br>(Min ; Max)<br>N valid | 24.00 (.)<br>(. ; .)<br>24.00 (24.00 ; 24.00)<br>(24.00 ; 24.00)<br>1 | . (.)<br>(. ; .)<br>. (. ; .)<br>(. ; .)<br>0 | 24.00 (.)<br>(. ; .)<br>24.00 (24.00 ; 24.00)<br>(24.00 ; 24.00)<br>1 | -- |
| **PATIENTS RECOVERED TO THE END OF THE OBSERVATIONAL PERIOD,** n (%)<br>Yes<br>No | **1 (100.0%)**<br>1 (100.0%)<br>0 (0.0%) | **--** | **1 (100.0%)**<br>1 (100.0%)<br>0 (0.0%) | **--** |
| **PATIENT RECOVERED- Days with some symptoms to the end of the observational period**<br>Mean (SD)<br>95%CI<br>Median (P25 ; P75)<br>(Min ; Max)<br>N valid | 24.00 (.)<br>(. ; .)<br>24.00 (24.00 ; 24.00)<br>(24.00 ; 24.00)<br>1 | -- | 24.00 (.)<br>(. ; .)<br>24.00 (24.00 ; 24.00)<br>(24.00 ; 24.00)<br>1 | -- |
| **9.15. Anorexia[3]** | | | | |

**Table S4. Recovery duration of each symptom associated with COVID-19 by symptoms**

| | Total sample | ImmunoFormulation cohort | Control cohort | p[1] |
|---|---|---|---|---|
| **Days with some symptoms to the end of the observational period** | | | | |
| Mean (SD) | 25.33 (1.15) | 26.00 (.) | 25.00 (1.41) | 1.0000 |
| 95%CI | (22.46 ; 28.20) | (. ; .) | (12.29 ; 37.71) | |
| Median (P25 ; P75) | 26.00 (24.00 ; 26.00) | 26.00 (26.00 ; 26.00) | 25.00 (24.00 ; 26.00) | |
| (Min ; Max) | (24.00 ; 26.00) | (26.00 ; 26.00) | (24.00 ; 26.00) | |
| N valid | 3 | 1 | 2 | |
| **PATIENTS RECOVERED TO THE END OF THE OBSERVATIONAL PERIOD,** n (%) | **3 (100.0%)** | **1 (100.0%)** | **2 (100.0%)** | **--** |
| Yes | 3 (100.0%) | 1 (100.0%) | 2 (100.0%) | |
| No | 0 (0.0%) | 0 (0.0%) | 0 (0.0%) | |
| **PATIENT RECOVERED-Days with some symptoms to the end of the observational period** | | | | |
| Mean (SD) | 25.33 (1.15) | 26.00 (.) | 25.00 (1.41) | 1.0000 |
| 95%CI | (22.46 ; 28.20) | (. ; .) | (12.29 ; 37.71) | |
| Median (P25 ; P75) | 26.00 (24.00 ; 26.00) | 26.00 (26.00 ; 26.00) | 25.00 (24.00 ; 26.00) | |
| (Min ; Max) | (24.00 ; 26.00) | (26.00 ; 26.00) | (24.00 ; 26.00) | |
| N valid | 3 | 1 | 2 | |
| **9.16. Myalgia[3]** | | | | |
| **Days with some symptoms to the end of the observational period** | | | | |
| Mean (SD) | 21.50 (12.02) | . (.) | 21.50 (12.02) | -- |
| 95%CI | (0.00 ; 129.50) | (. ; .) | (0.00 ; 129.50) | |
| Median (P25 ; P75) | 21.50 (13.00 ; 30.00) | . (. ; .) | 21.50 (13.00 ; 30.00) | |
| (Min ; Max) | (13.00 ; 30.00) | (. ; .) | (13.00 ; 30.00) | |
| N valid | 21.50 (12.02) | . (.) | 21.50 (12.02) | |
| **PATIENTS RECOVERED TO THE END OF THE OBSERVATIONAL PERIOD,** n (%) | **2 (100.0%)** | **--** | **2 (100.0%)** | **--** |
| Yes | 0 (0.0%) | | 0 (0.0%) | |
| No | 2 (100.0%) | | 2 (100.0%) | |
| **PATIENT RECOVERED-Days with some symptoms to the end of the observational period** | | | | |
| Mean (SD)<br>95%CI<br>Median (P25 ; P75)<br>(Min ; Max)<br>N valid | -- | -- | -- | -- |
| **9.17. Nasal congestion[3]** | | | | |
| **Days with some symptoms to the end of the observational period** | | | | |
| Mean (SD) | 30.00 (.) | . (.) | 30.00 (.) | -- |
| 95%CI | (. ; .) | (. ; .) | (. ; .) | |
| Median (P25 ; P75) | 30.00 (30.00 ; 30.00) | . (. ; .) | 30.00 (30.00 ; 30.00) | |
| (Min ; Max) | (30.00 ; 30.00) | (. ; .) | (30.00 ; 30.00) | |
| N valid | 1 | 0 | 1 | |

| Table S4. Recovery duration of each symptom associated with COVID-19 by symptoms | | | | |
|---|---|---|---|---|
| | Total sample | ImmunoFormulation cohort | Control cohort | p[1] |
| **PATIENTS RECOVERED TO THE END OF THE OBSERVATIONAL PERIOD,** n (%) | **1 (100.0%)** | **--** | **1 (100.0%)** | **--** |
| Yes | 0 (0.0%) | | 0 (0.0%) | |
| No | 1 (100.0%) | | 1 (100.0%) | |
| **PATIENT RECOVERED-Days with some symptoms to the end of the observational period** | | | | |
| Mean (SD) | -- | -- | -- | -- |
| 95%CI | | | | |
| Median (P25 ; P75) | | | | |
| (Min ; Max) | | | | |
| N valid | | | | |
| **9.18. Chest pain[3,4]** | | | | |
| **Days with some symptoms to the end of the observational period** | | | | |
| Mean (SD) | 6.00 (4.24) | 3.00 (.) | 9.00 (.) | -- |
| 95%CI | (0.00 ; 44.12) | (. ; .) | (. ; .) | |
| Median (P25 ; P75) | 6.00 (3.00 ; 9.00) | 3.00 (3.00 ; 3.00) | 9.00 (9.00 ; 9.00) | |
| (Min ; Max) | (3.00 ; 9.00) | (3.00 ; 3.00) | (9.00 ; 9.00) | |
| N valid | 2 | 1 | 1 | |
| **PATIENTS RECOVERED TO THE END OF THE OBSERVATIONAL PERIOD,** n (%) | **2 (100.0%)** | **1 (100.0%)** | **1 (100.0%)** | 1.0000(f) |
| Yes | 1 (50.0%) | 1 (100.0%) | 0 (0.0%) | |
| No | 1 (50.0%) | 0 (0.0%) | 1 (100.0%) | |
| **PATIENT RECOVERED-Days with some symptoms to the end of the observational period** | | | | |
| Mean (SD) | 3.00 (.) | 3.00 (.) | -- | -- |
| 95%CI | (. ; .) | (. ; .) | | |
| Median (P25 ; P75) | 3.00 (3.00 ; 3.00) | 3.00 (3.00 ; 3.00) | | |
| (Min ; Max) | (3.00 ; 3.00) | (3.00 ; 3.00) | | |
| N valid | 1 | 1 | | |
| **9.19. Hyporexia[3,4]** | | | | |
| **Days with some symptoms to the end of the observational period** | | | | |
| Mean (SD) | 16.50 (12.02) | 25.00 (.) | 8.00 (.) | -- |
| 95%CI | (0.00 ; 124.50) | (. ; .) | (. ; .) | |
| Median (P25 ; P75) | 16.50 (8.00 ; 25.00) | 25.00 (25.00 ; 25.00) | 8.00 (8.00 ; 8.00) | |
| (Min ; Max) | (8.00 ; 25.00) | (25.00 ; 25.00) | (8.00 ; 8.00) | |
| N valid | 2 | 1 | 1 | |
| **PATIENTS RECOVERED TO THE END OF THE OBSERVATIONAL PERIOD,** n (%) | **2 (100.0%)** | **1 (100.0%)** | **1 (100.0%)** | 1.0000(f) |
| Yes | 1 (50.0%) | 0 (0.0%) | 1 (100.0%) | |
| No | 1 (50.0%) | 1 (100.0%) | 0 (0.0%) | |
| **PATIENT RECOVERED-Days with some symptoms to the end of the observational period** | | | | |
| Mean (SD) | 8.00 (.) | -- | 8.00 (.) | -- |
| 95%CI | (. ; .) | | (. ; .) | |
| Median (P25 ; P75) | 8.00 (8.00 ; 8.00) | | 8.00 (8.00 ; 8.00) | |
| (Min ; Max) | (8.00 ; 8.00) | | (8.00 ; 8.00) | |
| N valid | 1 | | 1 | |

| Table S4. Recovery duration of each symptom associated with COVID-19 by symptoms | | | | |
|---|---|---|---|---|
| | **Total sample** | **ImmunoFormulation cohort** | **Control cohort** | p[1] |
| **9.20. Lymphedema[3,4]** | | | | |
| **Days with some symptoms to the end of the observational period** | | | | |
| Mean (SD) | 4.00 (.) | 4.00 (.) | . (.) | -- |
| 95%CI | (. ; .) | (. ; .) | (. ; .) | |
| Median (P25 ; P75) | 4.00 (4.00 ; 4.00) | 4.00 (4.00 ; 4.00) | . (. ; .) | |
| (Min ; Max) | (4.00 ; 4.00) | (4.00 ; 4.00) | (. ; .) | |
| N valid | 1 | 1 | 0 | |
| **PATIENTS RECOVERED TO THE END OF THE OBSERVATIONAL PERIOD,** n (%) | **1 (100.0%)** | **1 (100.0%)** | **--** | **--** |
| Yes | 1 (100.0%) | 1 (100.0%) | | |
| No | 0 (0.0%) | 0 (0.0%) | | |
| **PATIENT RECOVERED-Days with some symptoms to the end of the observational period** | | | | |
| Mean (SD) | 4.00 (.) | 4.00 (.) | -- | -- |
| 95%CI | (. ; .) | (. ; .) | | |
| Median (P25 ; P75) | 4.00 (4.00 ; 4.00) | 4.00 (4.00 ; 4.00) | | |
| (Min ; Max) | (4.00 ; 4.00) | (4.00 ; 4.00) | | |
| N valid | 1 | 1 | | |
| **9.21. Orthopnea[3,4]** | | | | |
| **Days with some symptoms to the end of the observational period** | | | | |
| Mean (SD) | 9.00 (.) | . (.) | 9.00 (.) | -- |
| 95%CI | (. ; .) | (. ; .) | (. ; .) | |
| Median (P25 ; P75) | 9.00 (9.00 ; 9.00) | . (. ; .) | 9.00 (9.00 ; 9.00) | |
| (Min ; Max) | (9.00 ; 9.00) | (. ; .) | (9.00 ; 9.00) | |
| N valid | 1 | 0 | 1 | |
| **PATIENTS RECOVERED TO THE END OF THE OBSERVATIONAL PERIOD,** n (%) | **1 (100.0%)** | **--** | **1 (100.0%)** | **--** |
| Yes | 0 (0.0%) | | 0 (0.0%) | |
| No | 1 (100.0%) | | 1 (100.0%) | |
| **PATIENT RECOVERED-Days with some symptoms to the end of the observational period** | | | | |
| Mean (SD) | -- | -- | -- | -- |
| 95%CI | | | | |
| Median (P25 ; P75) | | | | |
| (Min ; Max) | | | | |
| N valid | | | | |

[a] In patients who presented each symptom

[1] *Mann–Whitney U test or Fisher exact test (f)*

[2] *Variable generated by statistical programming.*

[3] *Other symptoms: According to MedDRA 23.0 (LLT)*

[4] *These symptoms were not first symptoms*

| Table S5. Recovery duration of each symptom associated with COVID-19 by symptoms | |
|---|---|
| | **ImmunoFormulation cohort** |
| **TOTAL RECOVERY FROM START OF IMMUNOFORMULATION TREATMENT[a]** | |
| **1. Fever** | |
| **Days with some symptoms to the end of the observational period** | |
| Mean (SD) | 2.25 (0.91) |
| 95%CI | (1.82 ; 2.68) |
| Median (P25 ; P75) | 2.00 (2.00 ; 3.00) |
| (Min ; Max) | (1.00 ; 5.00) |
| N valid | 20 |
| **PATIENTS RECOVERED TO THE END OF THE OBSERVATIONAL PERIOD,** n (%) | **20 (100.0%)** |
| Yes | 20 (100.0%) |
| No | 0 (0.0%) |
| **PATIENT RECOVERED- Days with some symptoms to the end of the observational period** | |
| Mean (SD) | 2.25 (0.91) |
| 95%CI | (1.82 ; 2.68) |
| Median (P25 ; P75) | 2.00 (2.00 ; 3.00) |
| (Min ; Max) | (1.00 ; 5.00) |
| N valid | 20 |
| **2. Dry Cough** | |
| **Days with some symptoms to the end of the observational period** | |
| Mean (SD) | 4.38 (6.31) |
| 95%CI | (0.57 ; 8.19) |
| Median (P25 ; P75) | 2.00 (2.00 ; 4.00) |
| (Min ; Max) | (1.00 ; 25.00) |
| N valid | 13 |
| **PATIENTS RECOVERED TO THE END OF THE OBSERVATIONAL PERIOD,** n (%) | **13 (100.0%)** |
| Yes | 13 (100.0%) |
| No | 0 (0.0%) |
| **PATIENT RECOVERED- Days with some symptoms to the end of the observational period** | |
| Mean (SD) | 4.38 (6.31) |
| 95%CI | (0.57 ; 8.19) |
| Median (P25 ; P75) | 2.00 (2.00 ; 4.00) |
| (Min ; Max) | (1.00 ; 25.00) |
| N valid | 13 |
| **3. Dyspnea** | |
| **Days with some symptoms to the end of the observational period** | |
| Mean (SD) | 3.67 (2.08) |
| 95%CI | (0.00 ; 8.84) |
| Median (P25 ; P75) | 3.00 (2.00 ; 6.00) |
| (Min ; Max) | (2.00 ; 6.00) |
| N valid | 3 |
| **PATIENTS RECOVERED TO THE END OF THE OBSERVATIONAL PERIOD,** n (%) | **3 (100.0%)** |
| Yes | 3 (100.0%) |
| No | 0 (0.0%) |
| **PATIENT RECOVERED- Days with some symptoms to the end of the observational period** | |
| Mean (SD) | 3.67 (2.08) |
| 95%CI | (0.00 ; 8.84) |
| Median (P25 ; P75) | 3.00 (2.00 ; 6.00) |
| (Min ; Max) | (2.00 ; 6.00) |
| N valid | 3 |
| **4. Loss of taste and smell** | |
| **Days with some symptoms to the end of the observational period** | |
| Mean (SD) | 19.73 (4.67) |

| Table S5. Recovery duration of each symptom associated with COVID-19 by symptoms | |
|---|---|
| | **ImmunoFormulation cohort** |
| 95%CI | (16.59 ; 22.87) |
| Median (P25 ; P75) | 20.00 (14.00 ; 23.00) |
| (Min ; Max) | (14.00 ; 26.00) |
| N valid | 11 |
| **PATIENTS RECOVERED TO THE END OF THE OBSERVATIONAL PERIOD,** n (%) | **11 (100.0%)** |
| Yes | 10 (90.9%) |
| No | 1 (9.1%) |
| **PATIENT RECOVERED- Days with some symptoms to the end of the observational period** | |
| Mean (SD) | 19.70 (4.92) |
| 95%CI | (16.18 ; 23.22) |
| Median (P25 ; P75) | 20.50 (14.00 ; 23.00) |
| (Min ; Max) | (14.00 ; 26.00) |
| N valid | 10 |
| **5. Headache** | |
| **Days with some symptoms to the end of the observational period** | |
| Mean (SD) | 2.00 (1.31) |
| 95%CI | (0.91 ; 3.09) |
| Median (P25 ; P75) | 2.00 (1.00 ; 2.00) |
| (Min ; Max) | (1.00 ; 5.00) |
| N valid | 8 |
| **PATIENTS RECOVERED TO THE END OF THE OBSERVATIONAL PERIOD,** n (%) | **8 (100.0%)** |
| Yes | 8 (100.0%) |
| No | 0 (0.0%) |
| **PATIENT RECOVERED- Days with some symptoms to the end of the observational period** | |
| Mean (SD) | 2.00 (1.31) |
| 95%CI | (0.91 ; 3.09) |
| Median (P25 ; P75) | 2.00 (1.00 ; 2.00) |
| (Min ; Max) | (1.00 ; 5.00) |
| N valid | 8 |
| **6. Diarrhea** | |
| **Days with some symptoms to the end of the observational period** | |
| Mean (SD) | 5.25 (5.85) |
| 95%CI | (0.00 ; 14.56) |
| Median (P25 ; P75) | 2.50 (2.00 ; 8.50) |
| (Min ; Max) | (2.00 ; 14.00) |
| N valid | 4 |
| **PATIENTS RECOVERED TO THE END OF THE OBSERVATIONAL PERIOD,** n (%) | **4 (100.0%)** |
| Yes | 4 (100.0%) |
| No | 0 (0.0%) |
| **PATIENT RECOVERED- Days with some symptoms to the end of the observational period** | |
| Mean (SD) | 5.25 (5.85) |
| 95%CI | (0.00 ; 14.56) |
| Median (P25 ; P75) | 2.50 (2.00 ; 8.50) |
| (Min ; Max) | (2.00 ; 14.00) |
| N valid | 4 |
| **7. Abdominal pain** | |
| **Days with some symptoms to the end of the observational period** | |
| Mean (SD) | 2.80 (1.30) |
| 95%CI | (1.18 ; 4.42) |
| Median (P25 ; P75) | 2.00 (2.00 ; 3.00) |
| (Min ; Max) | (2.00 ; 5.00) |
| N valid | 5 |
| **PATIENTS RECOVERED TO THE END OF THE OBSERVATIONAL PERIOD,** n (%) | **5 (100.0%)** |
| Yes | 5 (100.0%) |

| Table S5. Recovery duration of each symptom associated with COVID-19 by symptoms | |
|---|---|
| | **ImmunoFormulation cohort** |
| No | 0 (0.0%) |
| **PATIENT RECOVERED- Days with some symptoms to the end of the observational period** | |
| Mean (SD) | 2.80 (1.30) |
| 95%CI | (1.18 ; 4.42) |
| Median (P25 ; P75) | 2.00 (2.00 ; 3.00) |
| (Min ; Max) | (2.00 ; 5.00) |
| N valid | 5 |
| **8. Dermatological findings** | |
| **Days with some symptoms to the end of the observational period** | |
| Mean (SD) | 20.00 (.) |
| 95%CI | (. ; .) |
| Median (P25 ; P75) | 20.00 (20.00 ; 20.00) |
| (Min ; Max) | (20.00 ; 20.00) |
| N valid | 1 |
| **PATIENTS RECOVERED TO THE END OF THE OBSERVATIONAL PERIOD,** n (%) | **1 (100.0%)** |
| Yes | 0 (0.0%) |
| No | 1 (100.0%) |
| **PATIENT RECOVERED- Days with some symptoms to the end of the observational period** | **--** |
| Mean (SD) | |
| 95%CI | |
| Median (P25 ; P75) | |
| (Min ; Max) | |
| N valid | |
| **9.1. General discomfort[3]** | |
| **Days with some symptoms to the end of the observational period** | |
| Mean (SD) | 8.00 (8.16) |
| 95%CI | (0.00 ; 20.99) |
| Median (P25 ; P75) | 5.00 (3.00 ; 13.00) |
| (Min ; Max) | (2.00 ; 20.00) |
| N valid | 4 |
| **PATIENTS RECOVERED TO THE END OF THE OBSERVATIONAL PERIOD,** n (%) | **4 (100.0%)** |
| Yes | 3 (75.0%) |
| No | 1 (25.0%) |
| **PATIENT RECOVERED- Days with some symptoms to the end of the observational period** | |
| Mean (SD) | 4.00 (2.00) |
| 95%CI | (0.00 ; 8.97) |
| Median (P25 ; P75) | 4.00 (2.00 ; 6.00) |
| (Min ; Max) | (2.00 ; 6.00) |
| N valid | 3 |
| **9.2. Throat lesion[3]** | |
| **Days with some symptoms to the end of the observational period** | |
| Mean (SD) | 2.00 (.) |
| 95%CI | (. ; .) |
| Median (P25 ; P75) | 2.00 (2.00 ; 2.00) |
| (Min ; Max) | (2.00 ; 2.00) |
| N valid | 1 |
| **PATIENTS RECOVERED TO THE END OF THE OBSERVATIONAL PERIOD,** n (%) | **1 (100.0%)** |
| Yes | 1 (100.0%) |
| No | 0 (0.0%) |
| **PATIENT RECOVERED- Days with some symptoms to the end of the observational period** | |
| Mean (SD) | 2.00 (.) |
| 95%CI | (. ; .) |
| Median (P25 ; P75) | 2.00 (2.00 ; 2.00) |
| (Min ; Max) | (2.00 ; 2.00) |

| Table S5. Recovery duration of each symptom associated with COVID-19 by symptoms | |
|---|---|
| | **ImmunoFormulation cohort** |
| N valid | 1 |
| **9.3. Vomiting[3]** | |
| **Days with some symptoms to the end of the observational period** | |
| Mean (SD) | 2.00 (.) |
| 95%CI | (. ; .) |
| Median (P25 ; P75) | 2.00 (2.00 ; 2.00) |
| (Min ; Max) | (2.00 ; 2.00) |
| N valid | 1 |
| **PATIENTS RECOVERED TO THE END OF THE OBSERVATIONAL PERIOD,** n (%) | **1 (100.0%)** |
| Yes | 1 (100.0%) |
| No | 0 (0.0%) |
| **PATIENT RECOVERED- Days with some symptoms to the end of the observational period** | |
| Mean (SD) | 2.00 (.) |
| 95%CI | (. ; .) |
| Median (P25 ; P75) | 2.00 (2.00 ; 2.00) |
| (Min ; Max) | (2.00 ; 2.00) |
| N valid | 1 |
| **9.4. Weakness[3]** | |
| **Days with some symptoms to the end of the observational period** | |
| Mean (SD) | 1.92 (0.67) |
| 95%CI | (1.49 ; 2.34) |
| Median (P25 ; P75) | 2.00 (1.50 ; 2.00) |
| (Min ; Max) | (1.00 ; 3.00) |
| N valid | 12 |
| **PATIENTS RECOVERED TO THE END OF THE OBSERVATIONAL PERIOD,** n (%) | **12 (100.0%)** |
| Yes | 12 (100.0%) |
| No | 0 (0.0%) |
| **PATIENT RECOVERED- Days with some symptoms to the end of the observational period** | |
| Mean (SD) | 1.92 (0.67) |
| 95%CI | (1.49 ; 2.34) |
| Median (P25 ; P75) | 2.00 (1.50 ; 2.00) |
| (Min ; Max) | (1.00 ; 3.00) |
| N valid | 12 |
| **9.5. Sore throat[3]** | |
| **Days with some symptoms to the end of the observational period** | |
| Mean (SD) | 2.00 (.) |
| 95%CI | (. ; .) |
| Median (P25 ; P75) | 2.00 (2.00 ; 2.00) |
| (Min ; Max) | (2.00 ; 2.00) |
| N valid | 1 |
| **PATIENTS RECOVERED TO THE END OF THE OBSERVATIONAL PERIOD,** n (%) | **1 (100.0%)** |
| Yes | 1 (100.0%) |
| No | 0 (0.0%) |
| **PATIENT RECOVERED- Days with some symptoms to the end of the observational period** | |
| Mean (SD) | 2.00 (.) |
| 95%CI | (. ; .) |
| Median (P25 ; P75) | 2.00 (2.00 ; 2.00) |
| (Min ; Max) | (2.00 ; 2.00) |
| N valid | 1 |
| **9.6. Muscular pain[3]** | |
| **Days with some symptoms to the end of the observational period** | |
| Mean (SD) | 2.00 (.) |
| 95%CI | (. ; .) |
| Median (P25 ; P75) | 2.00 (2.00 ; 2.00) |

| Table S5. Recovery duration of each symptom associated with COVID-19 by symptoms | |
|---|---|
| | **ImmunoFormulation cohort** |
| (Min ; Max) | (2.00 ; 2.00) |
| N valid | 1 |
| **PATIENTS RECOVERED TO THE END OF THE OBSERVATIONAL PERIOD,** n (%) | **1 (100.0%)** |
| Yes | 1 (100.0%) |
| No | 0 (0.0%) |
| **PATIENT RECOVERED- Days with some symptoms to the end of the observational period** | |
| Mean (SD) | 2.00 (.) |
| 95%CI | (. ; .) |
| Median (P25 ; P75) | 2.00 (2.00 ; 2.00) |
| (Min ; Max) | (2.00 ; 2.00) |
| N valid | 1 |
| **9.7. Dehydration[3]** | |
| **Days with some symptoms to the end of the observational period** | |
| Mean (SD) | . (.) |
| 95%CI | (. ; .) |
| Median (P25 ; P75) | . (. ; .) |
| (Min ; Max) | (. ; .) |
| N valid | 0 |
| **PATIENTS RECOVERED TO THE END OF THE OBSERVATIONAL PERIOD,** n (%) | **--** |
| Yes | |
| No | |
| **PATIENT RECOVERED- Days with some symptoms to the end of the observational period** | |
| Mean (SD) | **--** |
| 95%CI | |
| Median (P25 ; P75) | |
| (Min ; Max) | |
| N valid | |
| **9.8. Emesis[3]** | |
| **Days with some symptoms to the end of the observational period** | |
| Mean (SD) | . (.) |
| 95%CI | (. ; .) |
| Median (P25 ; P75) | . (. ; .) |
| (Min ; Max) | (. ; .) |
| N valid | 0 |
| **PATIENTS RECOVERED TO THE END OF THE OBSERVATIONAL PERIOD,** n (%) | **--** |
| Yes | |
| No | |
| **PATIENT RECOVERED- Days with some symptoms to the end of the observational period** | **--** |
| Mean (SD) | |
| 95%CI | |
| Median (P25 ; P75) | |
| (Min ; Max) | |
| N valid | |
| **9.9. Hypoxemia[3]** | |
| **Days with some symptoms to the end of the observational period** | |
| Mean (SD) | . (.) |
| 95%CI | (. ; .) |
| Median (P25 ; P75) | . (. ; .) |
| (Min ; Max) | (. ; .) |
| N valid | 0 |
| **PATIENTS RECOVERED TO THE END OF THE OBSERVATIONAL PERIOD,** n (%) | **--** |
| Yes | |
| No | |

| Table S5. Recovery duration of each symptom associated with COVID-19 by symptoms | |
|---|---|
| | **ImmunoFormulation cohort** |
| **PATIENT RECOVERED- Days with some symptoms to the end of the observational period** | -- |
| Mean (SD) | |
| 95%CI | |
| Median (P25 ; P75) | |
| (Min ; Max) | |
| N valid | |
| **9.10. Dysuria[3]** | |
| **Days with some symptoms to the end of the observational period** | |
| Mean (SD) | . (.) |
| 95%CI | (. ; .) |
| Median (P25 ; P75) | . (. ; .) |
| (Min ; Max) | (. ; .) |
| N valid | 0 |
| **PATIENTS RECOVERED TO THE END OF THE OBSERVATIONAL PERIOD,** n (%) | -- |
| Yes | |
| No | |
| **PATIENT RECOVERED- Days with some symptoms to the end of the observational period** | -- |
| Mean (SD) | |
| 95%CI | |
| Median (P25 ; P75) | |
| (Min ; Max) | |
| N valid | |
| **9.11. Pollakiuria[3]** | |
| **Days with some symptoms to the end of the observational period** | |
| Mean (SD) | . (.) |
| 95%CI | (. ; .) |
| Median (P25 ; P75) | . (. ; .) |
| (Min ; Max) | (. ; .) |
| N valid | 0 |
| **PATIENTS RECOVERED TO THE END OF THE OBSERVATIONAL PERIOD,** n (%) | -- |
| Yes | |
| No | |
| **PATIENT RECOVERED- Days with some symptoms to the end of the observational period** | -- |
| Mean (SD) | |
| 95%CI | |
| Median (P25 ; P75) | |
| (Min ; Max) | |
| N valid | |
| **9.12. Sleepiness[3]** | |
| **Days with some symptoms to the end of the observational period** | |
| Mean (SD) | . (.) |
| 95%CI | (. ; .) |
| Median (P25 ; P75) | . (. ; .) |
| (Min ; Max) | (. ; .) |
| N valid | 0 |
| **PATIENTS RECOVERED TO THE END OF THE OBSERVATIONAL PERIOD,** n (%) | -- |
| Yes | |
| No | |
| **PATIENT RECOVERED- Days with some symptoms to the end of the observational period** | -- |
| Mean (SD) | |
| 95%CI | |
| Median (P25 ; P75) | |
| (Min ; Max) | |
| N valid | |

| Table S5. Recovery duration of each symptom associated with COVID-19 by symptoms | |
|---|---|
| | **ImmunoFormulation cohort** |
| **9.13. Apathy[3]** | |
| **Days with some symptoms to the end of the observational period** | |
| Mean (SD) | . (.) |
| 95%CI | (. ; .) |
| Median (P25 ; P75) | . (. ; .) |
| (Min ; Max) | (. ; .) |
| N valid | 0 |
| **PATIENTS RECOVERED TO THE END OF THE OBSERVATIONAL PERIOD,** n (%) | **--** |
| Yes | |
| No | |
| **PATIENT RECOVERED- Days with some symptoms to the end of the observational period** | **--** |
| Mean (SD) | |
| 95%CI | |
| Median (P25 ; P75) | |
| (Min ; Max) | |
| N valid | |
| **9.14. Disorientation[3]** | |
| **Days with some symptoms to the end of the observational period** | |
| Mean (SD) | . (.) |
| 95%CI | (. ; .) |
| Median (P25 ; P75) | . (. ; .) |
| (Min ; Max) | (. ; .) |
| N valid | 0 |
| **PATIENTS RECOVERED TO THE END OF THE OBSERVATIONAL PERIOD,** n (%) | **--** |
| Yes | |
| No | |
| **PATIENT RECOVERED- Days with some symptoms to the end of the observational period** | **--** |
| Mean (SD) | |
| 95%CI | |
| Median (P25 ; P75) | |
| (Min ; Max) | |
| N valid | |
| **9.15. Anorexia[3]** | |
| **Days with some symptoms to the end of the observational period** | |
| Mean (SD) | 26.00 (.) |
| 95%CI | (. ; .) |
| Median (P25 ; P75) | 26.00 (26.00 ; 26.00) |
| (Min ; Max) | (26.00 ; 26.00) |
| N valid | 1 |
| **PATIENTS RECOVERED TO THE END OF THE OBSERVATIONAL PERIOD,** n (%) | **1 (100.0%)** |
| Yes | 1 (100.0%) |
| No | 0 (0.0%) |
| **PATIENT RECOVERED- Days with some symptoms to the end of the observational period** | |
| Mean (SD) | 26.00 (.) |
| 95%CI | (. ; .) |
| Median (P25 ; P75) | 26.00 (26.00 ; 26.00) |
| (Min ; Max) | (26.00 ; 26.00) |
| N valid | 1 |
| **9.16. Myalgia[3]** | |
| **Days with some symptoms to the end of the observational period** | |
| Mean (SD) | . (.) |
| 95%CI | (. ; .) |
| Median (P25 ; P75) | . (. ; .) |
| (Min ; Max) | (. ; .) |

| Table S5. Recovery duration of each symptom associated with COVID-19 by symptoms | |
|---|---|
| | **ImmunoFormulation cohort** |
| N valid | 0 |
| **PATIENTS RECOVERED TO THE END OF THE OBSERVATIONAL PERIOD,** n (%) | **--** |
| Yes | |
| No | |
| **PATIENT RECOVERED- Days with some symptoms to the end of the observational period** | **--** |
| Mean (SD) | |
| 95%CI | |
| Median (P25 ; P75) | |
| (Min ; Max) | |
| N valid | |
| **9.17. Nasal congestion[3]** | |
| **Days with some symptoms to the end of the observational period** | |
| Mean (SD) | . (.) |
| 95%CI | (. ; .) |
| Median (P25 ; P75) | . (. ; .) |
| (Min ; Max) | (. ; .) |
| N valid | 0 |
| **PATIENTS RECOVERED TO THE END OF THE OBSERVATIONAL PERIOD,** n (%) | **--** |
| Yes | |
| No | |
| **PATIENT RECOVERED- Days with some symptoms to the end of the observational period** | **--** |
| Mean (SD) | |
| 95%CI | |
| Median (P25 ; P75) | |
| (Min ; Max) | |
| N valid | |
| **9.18. Chest pain[3,4]** | |
| **Days with some symptoms to the end of the observational period** | |
| Mean (SD) | 3.00 (.) |
| 95%CI | (. ; .) |
| Median (P25 ; P75) | 3.00 (3.00 ; 3.00) |
| (Min ; Max) | (3.00 ; 3.00) |
| N valid | 1 |
| **PATIENTS RECOVERED TO THE END OF THE OBSERVATIONAL PERIOD,** n (%) | **1 (100.0%)** |
| Yes | 1 (100.0%) |
| No | 0 (0.0%) |
| **PATIENT RECOVERED- Days with some symptoms to the end of the observational period** | |
| Mean (SD) | 3.00 (.) |
| 95%CI | (. ; .) |
| Median (P25 ; P75) | 3.00 (3.00 ; 3.00) |
| (Min ; Max) | (3.00 ; 3.00) |
| N valid | 1 |
| **9.19. Hyporexia[3,4]** | |
| **Days with some symptoms to the end of the observational period** | |
| Mean (SD) | 24.00 (.) |
| 95%CI | (. ; .) |
| Median (P25 ; P75) | 24.00 (24.00 ; 24.00) |
| (Min ; Max) | (24.00 ; 24.00) |
| N valid | 1 |
| **PATIENTS RECOVERED TO THE END OF THE OBSERVATIONAL PERIOD,** n (%) | **1 (100.0%)** |
| Yes | 0 (0.0%) |
| No | 1 (100.0%) |
| **PATIENT RECOVERED- Days with some symptoms to the end of the observational period** | **--** |

| Table S5. Recovery duration of each symptom associated with COVID-19 by symptoms | |
|---|---|
| | **ImmunoFormulation cohort** |
| Mean (SD)<br>95%CI<br>Median (P25 ; P75)<br>(Min ; Max)<br>N valid | |
| **9.20. Lymphedema[3,4]** | |
| **Days with some symptoms to the end of the observational period** | |
| Mean (SD) | 4.00 (.) |
| 95%CI | (. ; .) |
| Median (P25 ; P75) | 4.00 (4.00 ; 4.00) |
| (Min ; Max) | (4.00 ; 4.00) |
| N valid | 1 |
| **PATIENTS RECOVERED TO THE END OF THE OBSERVATIONAL PERIOD,** n (%) | **1 (100.0%)** |
| Yes | 1 (100.0%) |
| No | 0 (0.0%) |
| **PATIENT RECOVERED- Days with some symptoms to the end of the observational period** | |
| Mean (SD) | 4.00 (.) |
| 95%CI | (. ; .) |
| Median (P25 ; P75) | 4.00 (4.00 ; 4.00) |
| (Min ; Max) | (4.00 ; 4.00) |
| N valid | 1 |
| **9.21. Orthopnea[3,4]** | |
| **Days with some symptoms to the end of the observational period** | |
| Mean (SD) | . (.) |
| 95%CI | (. ; .) |
| Median (P25 ; P75) | . (. ; .) |
| (Min ; Max) | (. ; .) |
| N valid | 0 |
| **PATIENTS RECOVERED TO THE END OF THE OBSERVATIONAL PERIOD,** n (%) | **--** |
| Yes | |
| No | |
| **PATIENT RECOVERED- Days with some symptoms to the end of the observational period** | **--** |
| Mean (SD) | |
| 95%CI | |
| Median (P25 ; P75) | |
| (Min ; Max) | |
| N valid | |

[a] In patients who presented each symptom

[1] *Mann–Whitney U test*

[2] *Variable generated by statistical programming.*

[3] *Other symptoms: According to MedDRA 23.0 (LLT)*

[4] *These symptoms were not first symptoms*
